# Effects of family planning counseling delivered during maternal healthcare on postpartum modern contraceptive uptake in low- and middle-income countries: a systematic review and meta-analysis

**DOI:** 10.1101/2024.09.29.24314560

**Authors:** Md Nuruzzaman Khan, Atika Rahman Chowdhury, Md Shohel Rana, Rashed Hossain, Tahir Ahmed Hassen, Catherine Chojenta, Melissa L. Harris

**Author notes:** **Corresponding author:** Md Nuruzzaman Khan, PhD, Associate Professor, Department of Population Science, Jatiya Kabi Kazi Nazrul Islam University, Mymensingh, Bangladesh.

## Abstract

**Background:** Postpartum modern contraception is crucial for preventing unintended pregnancies, reducing short inter-pregnancy intervals, and improving maternal and child health outcomes. Family planning counseling, when engaging with maternal healthcare services, may enhance modern contraceptive uptake in the postpartum period. However, evidence in low- and middle-income countries (LMICs) has yielded conflicting findings. We aimed to explore the effects of family planning counseling delivered as part of maternal healthcare on postpartum modern contraceptive uptake in LMICs.

**Methods:** The following six databases were searched in April 2024: PubMed, Web of Science, Embase, Global Health, Medline, and Scopus. Studies that examined the association between family planning counseling and modern contraception uptake in the postpartum period were included. The findings of the included studies were summarized narratively and through a random-effects meta-analysis when data supported. Trim and fill estimates and subgroup analyses were conducted to address publication bias and heterogeneity, respectively.

**Results:** A total of 61 studies were included, of which 42 studies were considered for quantitative synthesis and the remaining 19 studies included in a narrative synthesis. We found that women who received family planning counseling during maternal healthcare visits were 2.75 times (95% CI, 2.11-3.58) more likely to use modern contraception during the postpartum period compared to those who did not receive family planning counseling. Although we observed significant publication bias and heterogeneity, addressing these issues through trim and fill estimation and subgroup analyses, respectively, produced estimates consistent with the summary findings. A narrative synthesis of an additional 19 studies also supports the effectiveness of family planning counseling during maternal healthcare visits on postpartum modern contraception uptake.

**Conclusion:** Integrating family planning counseling into maternal healthcare can significantly increase modern contraceptive use in the postpartum period, as such effectively reducing unintended and short interval pregnancy as well as adverse maternal and child health outcomes. Policymakers should prioritize training healthcare providers and developing standardized protocols for effective counseling.

**Research in context:** *Evidence Before the Study:* Postpartum contraception is essential for improving maternal and child health, but evidence from LMICs on the impact of family planning counselling during maternal healthcare on contraceptive uptake has been mixed. While some studies show a positive association, others do not, and no comprehensive meta-analysis had been conducted on this topic. This highlights a critical research gap, emphasizing the need for a thorough study to resolve these conflicting findings and inform policy and program development.

*Added Value of the Study:* Our study is the largest to date, synthesizing evidence from 61 studies across LMICs. We found that women receiving family planning counselling during maternal healthcare visits were 2.75 times more likely to use modern contraception postpartum. We addressed publication bias and heterogeneity, providing robust evidence of the intervention’s effectiveness.

*Implication of the Study Findings:* Our findings highlight the importance of integrating family planning counselling into maternal healthcare services in LMICs. Policymakers should invest in training healthcare providers and standardizing protocols to improve postpartum contraceptive uptake, reduce unintended pregnancies, and enhance maternal and child health outcomes.

## Introduction

With approximately two-thirds of the time allotted for achieving the Sustainable Development Goals (SDGs) between 2015 and 2030 already elapsed, the global community, particularly in low- and middle-income countries (LMICs), is actively assessing its progress concerning maternal and child health. Of particular concern in this evaluation is the high prevalence of unintended pregnancies (47%) and short birth intervals (23%), both of which significantly contribute to adverse health outcomes such as unsafe abortions, pregnancy complications, and limited access to maternal healthcare services ^1,2^. The long-term ramifications of unintended pregnancies and short birth intervals are also evident, including malnutrition, increased likelihood of school dropout, and perpetuating cycles of disadvantage ^1,3-5^. This issue becomes more acute when pregnancies occur shortly after a previous birth, as the mother’s body may not be adequately prepared, and she may lack the physical and financial resources to support two young children simultaneously ^1,4,6^. Thus, the combined occurrence of unintended pregnancies and short birth intervals poses significant challenges to achieving the SDGs’ core target of “leaving no one behind”. They can also influence specific targets related to healthcare access and outcomes, including universal access to maternal healthcare services (target 3.8), reduction of maternal mortality to 70 per 100,000 live births (target 3.1), and reduction of neonatal and child mortality to 12 and 25 per 1,000 live births, respectively (target 3.2). Addressing these issues is crucial for improving maternal and child health and achieving the SDGs.

Around half of all unintended pregnancies and short birth intervals occur during the postpartum period, spanning from childbirth to 12 months ^7^. The primary reasons for this include non-use of effective contraception or reliance on less effective methods ^8,9^. Several misconceptions at the community level further exacerbate this issue, such as the belief that women are infertile during this time or that breastfeeding prevents conception ^10,11^. Additionally, concerns about the impact of contraception, particularly hormonal contraception, on breastfeeding and newborns deter some women from utilizing contraception during this critical period ^10,12^. These misconceptions are particularly prevalent among disadvantaged groups, including those who are illiterate and impoverished, among whom unintended pregnancies and short birth intervals are even more common ^1,13^. Importantly, the postpartum period presents a critical opportunity for initiating effective contraception, as a substantial number of women are highly motivated to prevent unintended pregnancies and plan future births during this time ^14,15^. Recognizing this, the World Health Organization (WHO) has advocated for scaling up the use of modern contraception during the postpartum period, and this issue has been prioritized as a key indicator of national Family Planning programs in LMICs ^16^.

Ensuring informed decision-making about contraception after childbirth requires comprehensive postpartum contraceptive counseling during any time point during maternal healthcare access ^17-19^. This counseling provides women with detailed information about family planning and various contraceptive methods, including their efficacy in preventing pregnancy, safety, and suitability while breastfeeding ^20,21^. It also addresses misconceptions and concerns regarding hormonal contraceptives and their potential impact on breast milk supply ^21,22^. As such, health policies in LMICs view postpartum contraceptive counseling as a key strategy for increasing the uptake of modern contraception(e.g. hormonal pills, implants, intrauterine devices (IUDs), condoms, sterilization, and injectable contraceptives) during this period where LARCs are the most effective in preventing unintended pregnancy and SBIs but uptake is low ^5,23,24^. However, systematic assessments of the effectiveness of postpartum counseling on contraception uptake in LMICs are lacking, with few studies conducted, primarily in African countries, and inconclusive findings identified from those

conducted in other regions, including Asia ^25-27^. While researchers from Africa, particularly in Ethiopia, have attempted to reconcile these disparities through systematic reviews and meta-analyses, such efforts have been limited to specific countries or regions within Africa ^25-28^. As a result, the true effects of postpartum contraception counseling on modern contraception uptake during the postpartum period in LMICs remain unknown. We therefore conducted this study to explore the effects of postpartum counseling on modern contraception uptake in the postpartum period in LMICs.

## Methods

We performed a systematic review and meta-analysis in adherence with the Preferred Reporting Items for Systematic Reviews and Meta-Analyses (PRISMA) guidelines for observational studies. This approach ensured a comprehensive and transparent evaluation of the relationship between contraceptive counseling and postpartum modern contraception uptake.

### Search strategy

A systematic literature search was conducted in April 2024 in six databases: PubMed, Web of Science, Embase, Global Health, Medline, and Scopus. Studies published since the establishment of the SDGs in January 2015 to April 2024 were included. Searches were based on individual comprehensive search strategies for each database. We developed search strategies using a combination of free text words, words in titles/abstracts, and Medical Subject Headings (MeSH) terms related to exposure, outcomes, and settings. The exposure-related terms were family planning counseling, family planning information, family planning advice, or contraceptive counseling during any stage of maternal healthcare service provision, including antenatal, delivery, and postnatal healthcare services. The outcomes-related terms were modern contraception uptake during the postpartum period, including contraception or modern contraception or name of modern contraception. We also composed country restrictions and included LMICs (classified based on gross national income (GNI) per capita, calculated using the World Bank Atlas method and encompassed low-income countries (GNI $1085 or less in 2021) and lower middle income countries (GNI $1086 to $4255) ^29^. The term LMICs was also considered together with country names. These terms were combined using the Boolean operators (AND, OR). The full search strategy and search results for each database are presented in Supplementary Tables 01 to 06. Further searches for eligible studies were conducted by reviewing selected journals’ websites and reference lists of the selected articles.

### Study eligibility criteria

Eligible articles had to meet the following criteria: (i) peer-reviewed journal articles, (ii) written in English, (iii) published from 2015 onwards following the initiation of SDGs, (iv) reported on family planning counseling at any stage of maternal healthcare services access, including antenatal, delivery, and postnatal healthcare services, (v) reported information on postpartum contraception uptake, and (vi) conducted in any LMICs.

### Exclusion criteria

We excluded studies involving (i) reproductive-aged women with high-risk characteristics such as HIV/AIDS or cancer as they may require targeted counselling compared to the general population, (ii) contraception use outside the postpartum period, (iii) counseling on issues other than family planning or where family planning counseling could not be disaggregated for modern contraception uptake, (iv) studies conducted outside LMICs, and (v) studies published before 2015. Commentaries, letters to the editor, conference papers, and these were also excluded.

### Study selection

Two authors (ARC and MSR) independently reviewed all articles, starting with title and abstract screening. Articles selected at this stage were considered for full-text review, which was conducted by two authors (ARC and RH). Disagreements were resolved through discussion, involving the lead author (MNK) if necessary. Online platforms such as COVIDENCE, EndNote 21, and face-to-face meetings were used to complete this review.

### Data extraction

Prior to tabulating the final dataset, a data extraction template was designed, trialed, and modified following the “Strengthening the Reporting of Observational Studies in Epidemiology” guidelines. Two authors (ARC and MRH) independently extracted relevant data, including authors’ names, study design, sample size, study setting, type of maternal healthcare services accessed when counseling was received, types of counseling received, and postpartum contraceptive uptake. Reported effect sizes (odds ratios, ORs), underlying data used to calculate ORs, and whether they were adjusted or unadjusted for possible confounders were also extracted. Disagreements between the data extractors were resolved through discussion, involving the lead author (MNK) if necessary. Some articles included in this review used slightly different intervals to define the postpartum period. However, for the quantitative synthesis, we followed the WHO classification of postpartum period (1 to 12 months after delivery) ^30^. Articles that did not follow the WHO classification were synthesized narratively.

### Quality assessment in included studies

We used the modified Newcastle-Ottawa Scale (NOS) to assess the quality of the included studies ^31^. The items included in the scale differed for cross-sectional, case-control, cohort studies, and randomized control trials. Two authors (ARC and RH) assessed included articles, assigning 1 point for each item if the study met the relevant item condition. Aggregated scores were used to measure overall study quality as good (score: 8 to 9), moderate (score: 5 to 7), and low (score: <5).

### Exposure variable

The primary exposure of interest was family planning counseling that women received during maternal healthcare visits, including antenatal, delivery, and postnatal healthcare services. Responses were extracted dichotomously as “Yes” if women reported counseling at any stage of maternal healthcare services access and “No” if they reported no counseling at any stage. If women reported receiving counseling at more than one stage, their data were extracted separately for each instance. Outcome variable

The outcome variable considered was uptake of modern contraception during the postpartum period. We considered the WHO classification of the postpartum period, for inclusion in the quantitative synthesis. Studies that did not strictly follow this classification to define the postpartum period were summarized narratively.

### Statistical analysis

We used extracted odds ratio (ORs) as the basis of analysis. If ORs were unavailable in the paper, we calculated unadjusted ORs. Fixed-effects and random-effects models were used to calculate the summary estimate for the overall effects of family planning counseling on modern contraception uptake during the postpartum period. The final model was selected based on the heterogeneity assessment using the I^2^ statistic with a p-value. According to this statistic, the random-effects model is applicable when heterogeneity (I^2^ statistic) is moderate (50-74%) or high (75-100%). This was met in our study findings; thus, the random-effects model was used to summarize the findings. We also explored the source of heterogeneity using subgroup analysis and meta-regression once moderate or higher heterogeneity was identified. Pre-specified subgroups included study sample size, confounding adjustment, study design, study settings, and the number of stages during maternal healthcare services when family planning counseling was received. Publication bias was explored by visual inspection of Funnel Plot asymmetry and Egger’s regression test. The trim-and-fill method was used when evidence of publication bias was found, and to estimate and adjust for potentially missing studies, the effect size was recalculated accordingly. Statistical software, STATA version 15.1 (Stata Corp), was used for all analyses.

## Results

### Search results

We identified a total of 3,178 articles from six databases, including 90 additional articles obtained through searches of relevant journal websites, Google, Google Scholar, and reference lists of selected articles. After removing 523 duplicate studies, 2,655 unique articles remained. From these, 2,351 studies were excluded after screening the titles and abstracts. We then conducted full-text reviews on the remaining 111 articles, excluding 50 based on the full-text review. Ultimately, 61 studies were included in our analysis: 42 in the quantitative and 19 in the narrative synthesis.

#### Study characteristics

The total sample analyzed in the 61 included studies was 1,55,552. The majority of the studies were conducted in Ethiopia (n=34), followed by Nepal (n=6), Uganda (n=4), India (n=3), Ghana (n=3), Tanzania (n=3), and Indonesia (n=2) (Supplementary Table 07). Of these studies, 41 employed a cross-sectional design. More than two-thirds (n=41) analyzed facility-level data, and 40 studies reported adjusted odds ratios (aORs). Most studies indicated that participants received family planning counseling during antenatal healthcare only (n=30), followed by those who received counseling during both antenatal healthcare and postnatal healthcare (n=13). All studies included were moderate to good quality (Supplementary Table 08-10).

#### Effect of family planning counseling during maternal healthcare services on modern contraception uptake in the postpartum period

We determined the summary effects of family planning counseling on modern contraception uptake during the postpartum period through a random effects meta-analysis. We found that the counseled group was 2.75 times (95% CI, 2.11-3.58) more likely to report the use of modern contraception during the postpartum period compared to the non-counseled group (Figure 2). However, there was significant evidence of publication bias (Figure 3a). After adjusting for this bias using the trim-and-fill method, we identified 12 potentially missing studies (Figure 3(b)). Including these in the analysis indicated that those who received counseling were 1.93 times (95% CI, 1.48-2.51) more likely to use modern contraception during the postpartum period compared to those who did not receive counseling (results not shown in the table or figure).

**Figure 1:**
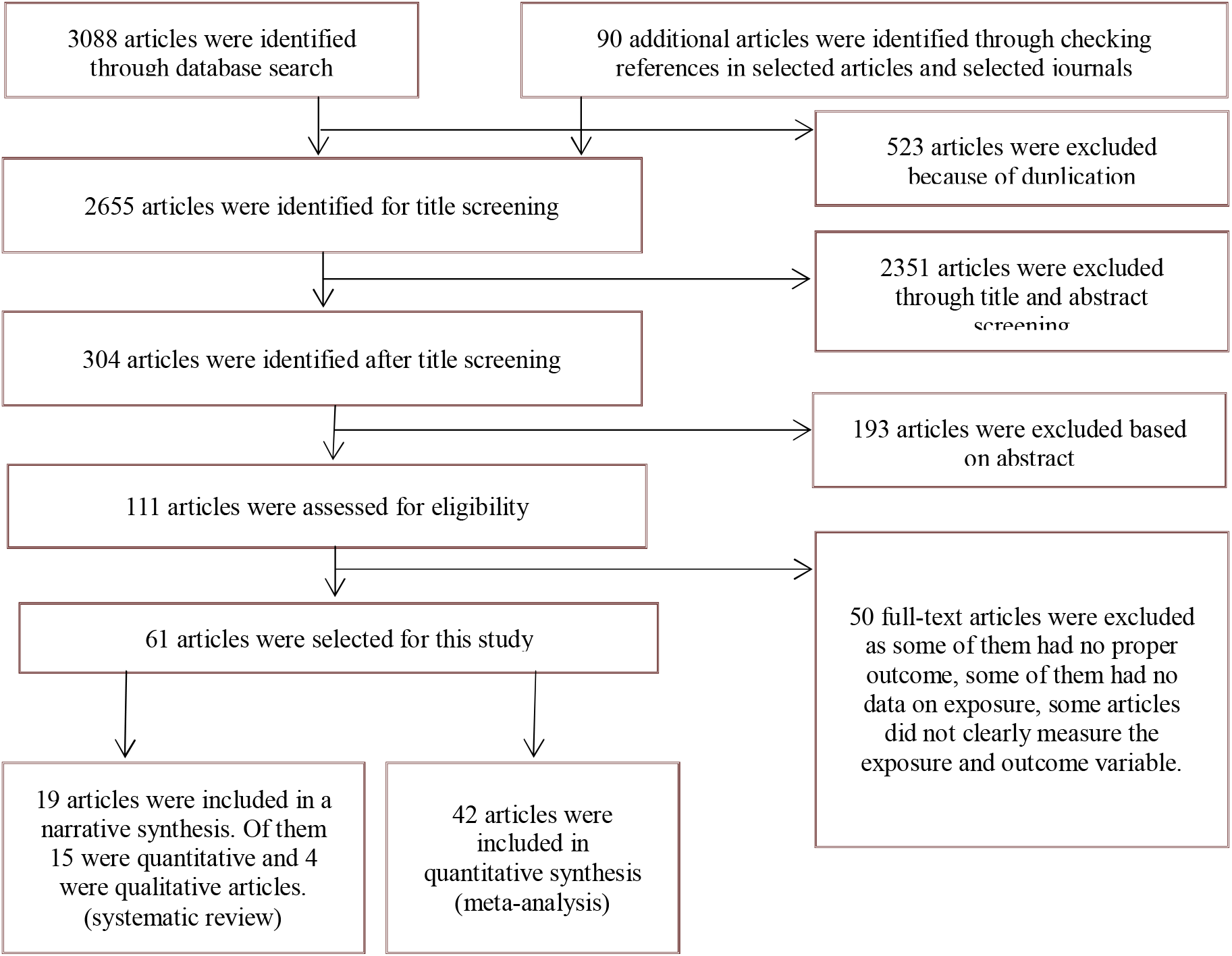
Schematic presentation of the studies included in the systematic review

**Figure 2:**
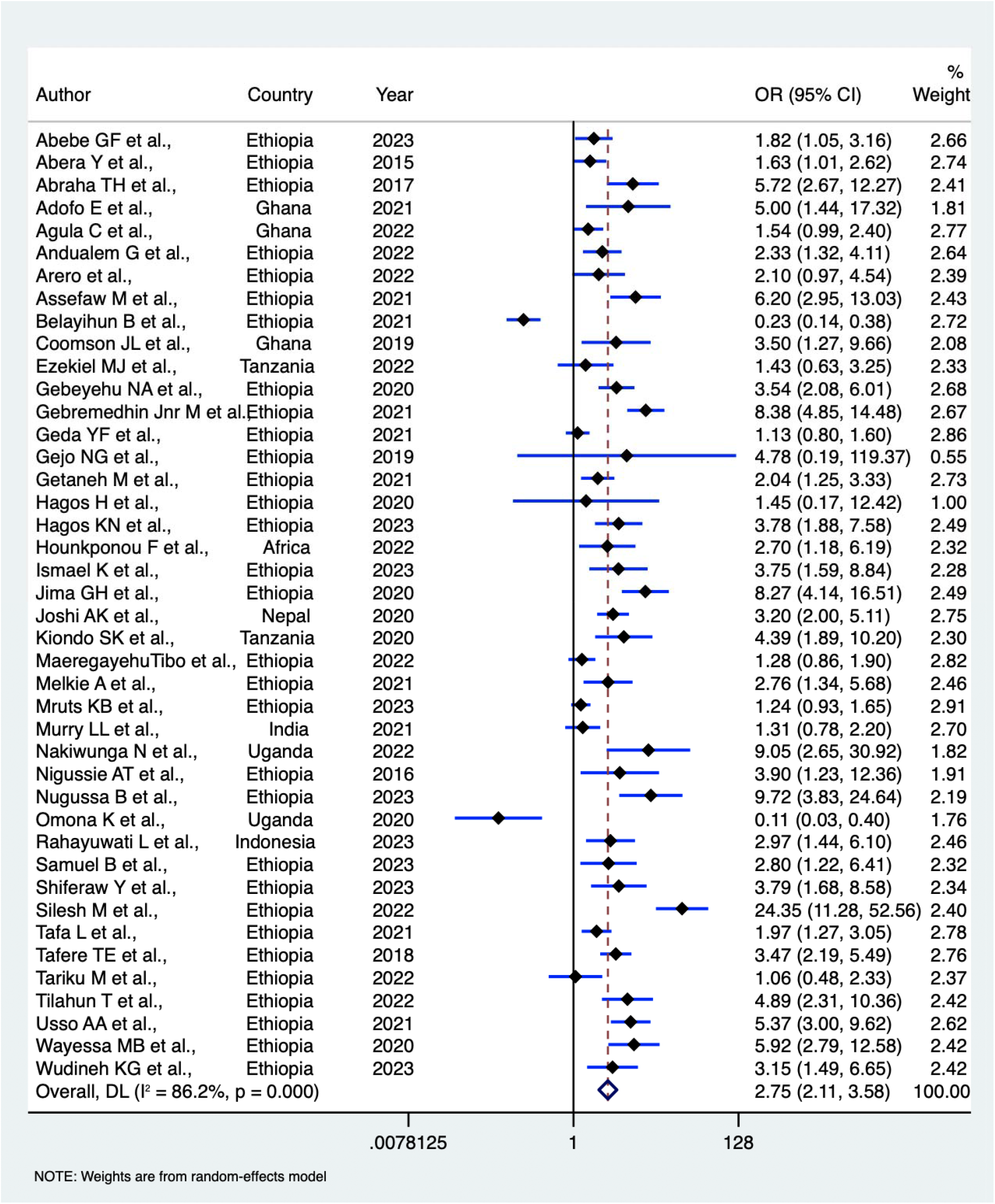
Summary effects of counselling received during accessing maternal healthcare services on modern contraception uptake during postpartum period.

**Figure 3:**
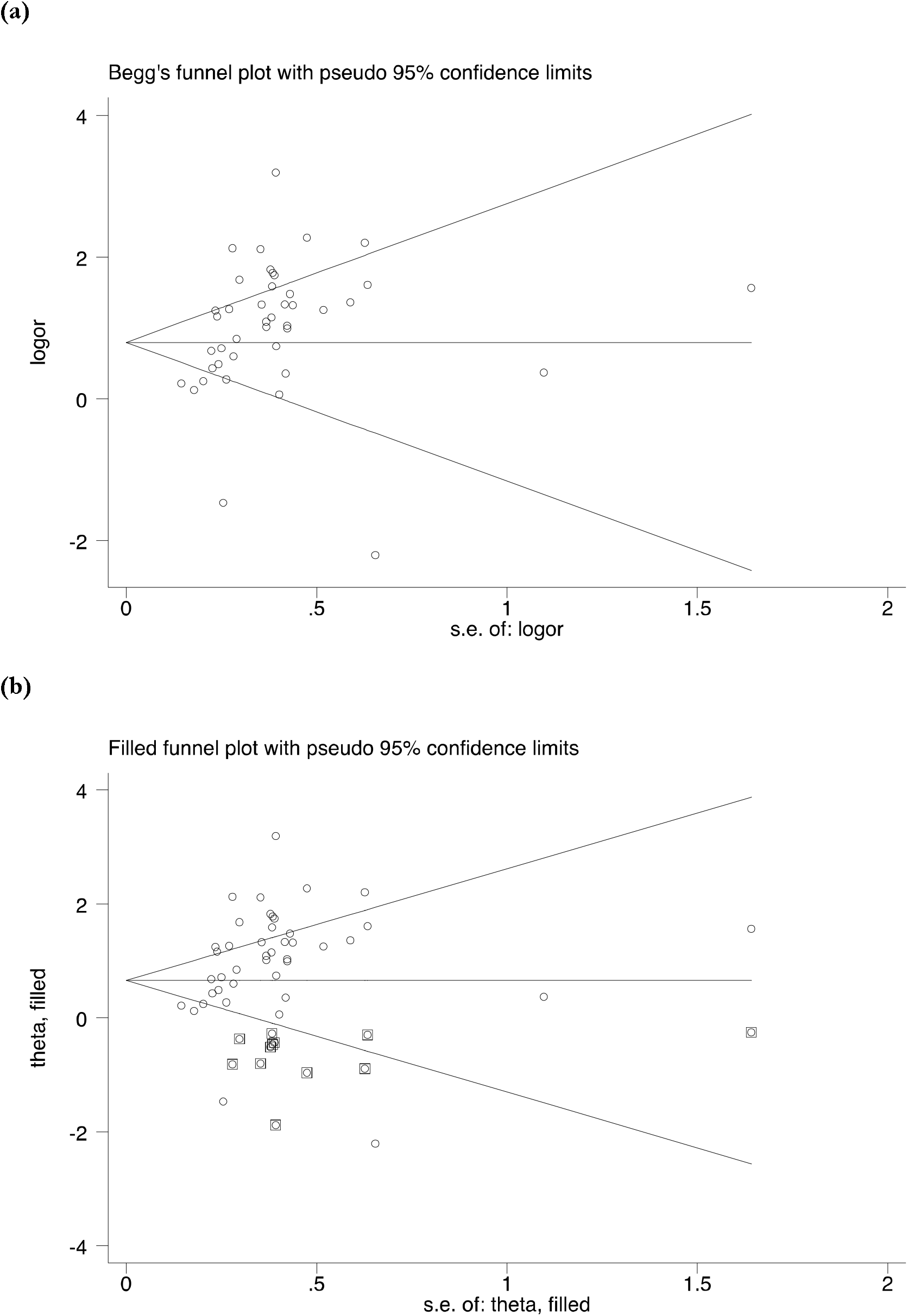
Funnel plots to access publication bias (a) and addressing publication bias through trim and fill estimate (b).

#### Stratified analysis

We found evidence of moderate to high heterogeneity in the likelihood of modern contraception uptake during the postpartum period between the counseled group and the non-counseled group. To investigate the sources of this heterogeneity, we conducted a stratified analysis based on various characteristics, including sample size, study setting, confounding factors adjustment, study design, timing of contraceptive counseling, and country of study (Table 1, Supplementary figure 01-06). While the likelihood of modern contraception uptake varied across these characteristics, the direction of the effect remained consistent with the pooled estimates in all cases.

**Table 1:** Stratified analysis of pooled Odd Ratio (OR) of modern contraception uptake in the postpartum period across selected characteristics

|  | Pooled Odds Ratio (OR)<br>(95% CI) | Heterogeneity, *p* | Meta-regression, *p* |
| --- | --- | --- | --- |
| **Sample size** |  |  |  |
| ≤490 | 2.96 (2.14-4.09) | <0.01 | 0.494 |
| >490 | 2.46 (1.59-3.80) | <0.01 |  |
| **Study setting** |  |  |  |
| Facility based study | 2.80 (1.83-4.29) | <0.01 | 0.619 |
| Community based study | 3.02 (2.16-4.24) | <0.01 |  |
| Regional level study | 1.80 (1.19-1.73) | <0.05 |  |
| National level study | 1.24 (0.93-1.65)<sup>+</sup> | NA |  |
| **Confounding factors** |  |  |  |
| Adjusted | 2.69 (2.04-3.56) | <0.01 | 0.745 |
| Unadjusted | 2.30 (1.06-5.01) | 0.094 |  |
| **Study design** |  |  |  |
| Cross-sectional study | 2.73 (2.01-3.69) | <0.01 |  |
| Case-control study | 4.76 (2.87-7.92) | 0.340 | 0.589 |
| Cohort (prospective or retrospective) study | 2.25 (1.18-4.27) | <0.01 |  |
| **Time of contraceptive counseling received<sup>++</sup>** |  |  |  |
| During ANC only | 3.01 (1.94-4.69) | <0.01 | 0.712 |
| During ANC and DHC | 3.90 (1.23-12.36)<sup>+</sup> | NA |  |
| During ANC and PNC | 2.35 (1.75-3.17) | <0.01 |  |
| During ANC, DHC and PNC | 1.97 (1.27-3.05)<sup>+</sup> | NA |  |
| **Country of the study** |  |  |  |
| African region | 2.70 (1.18-6.19)<sup>+</sup> | NA | 0.400 |
| Ethiopia | 2.96 (2.15-4.07) | <0.01 |  |
| Ghana | 2.54 (1.19-5.41) | <0.01 |  |
| India | 1.31 (0.78-2.20)<sup>+</sup> | NA |  |
| Indonesia | 2.97 (1.44-6.10)<sup>+</sup> | NA |  |
| Nepal | 3.20 (2.00-5.11)<sup>+</sup> | NA |  |
| Tanzania | 2.49 (0.83-7.49) | <0.05 |  |
| Uganda | 1.00 (0.001-75.45) | <0.01 |  |

Note: \*Odds ratio for single study, Meta regression p values represent a test of the entire characteristics, \*\*ANC, Antenatal care, \*\*DHC, Delivery healthcare, \*\*PNC, Postnatal healthcare.

**Table 2:**
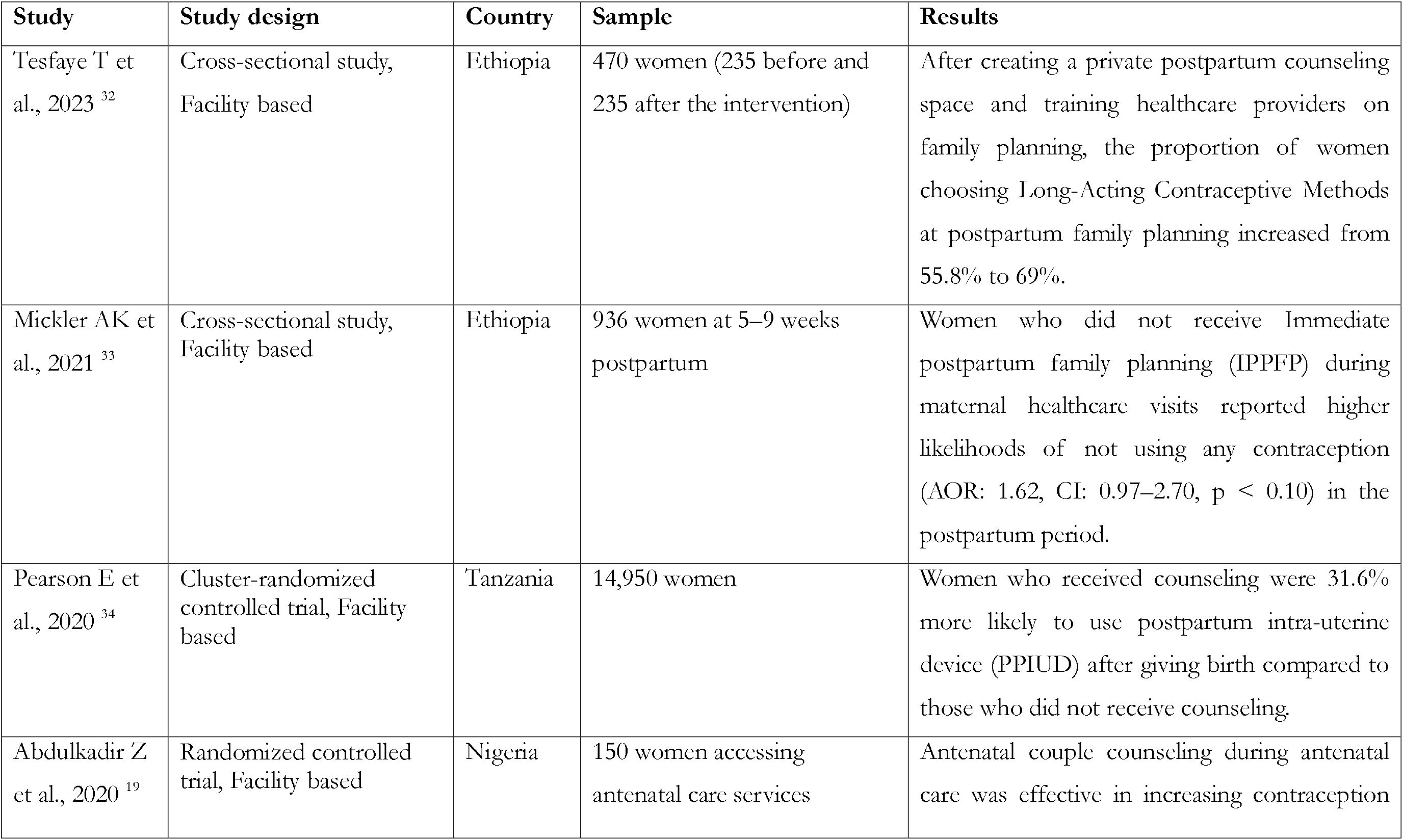
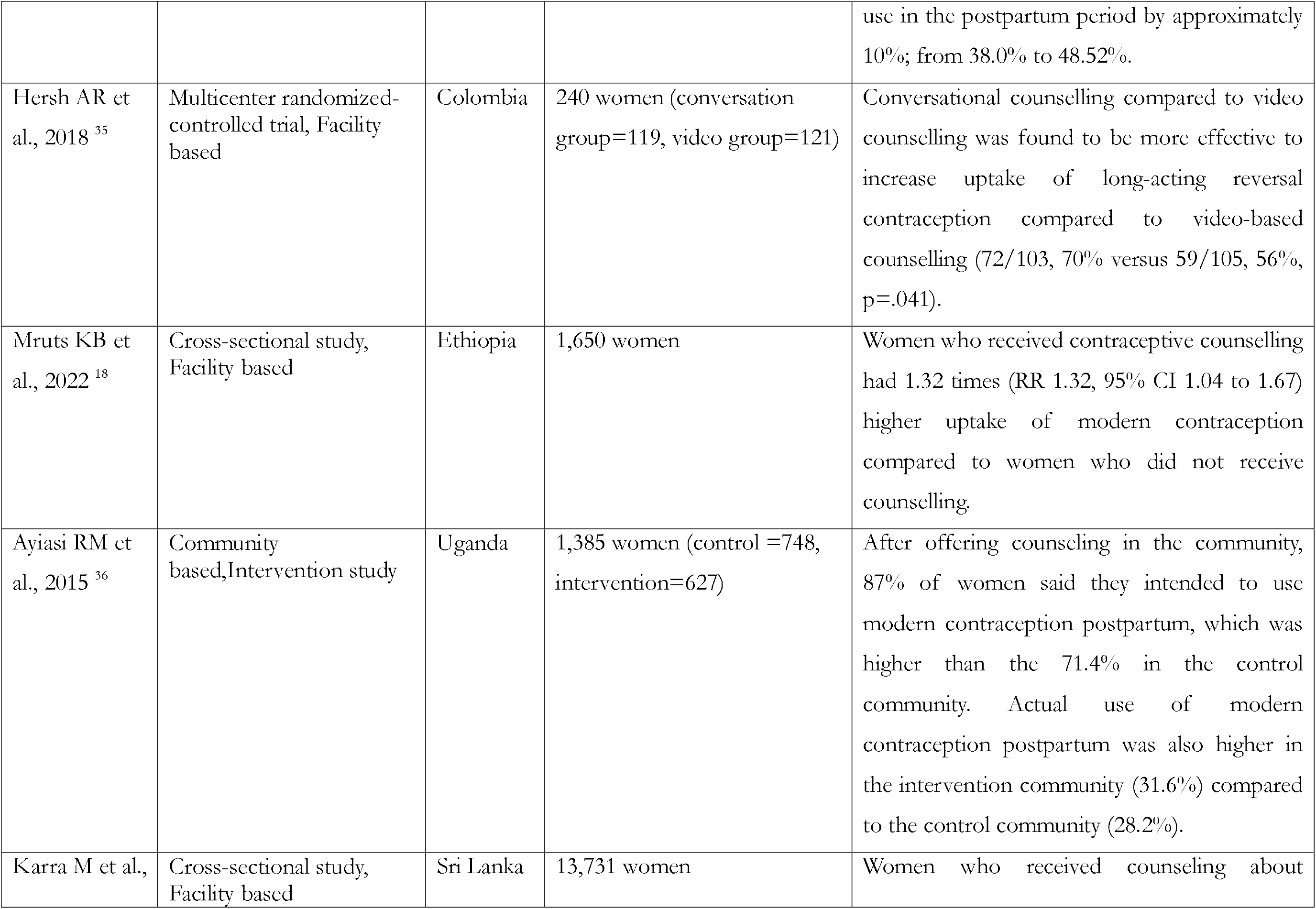
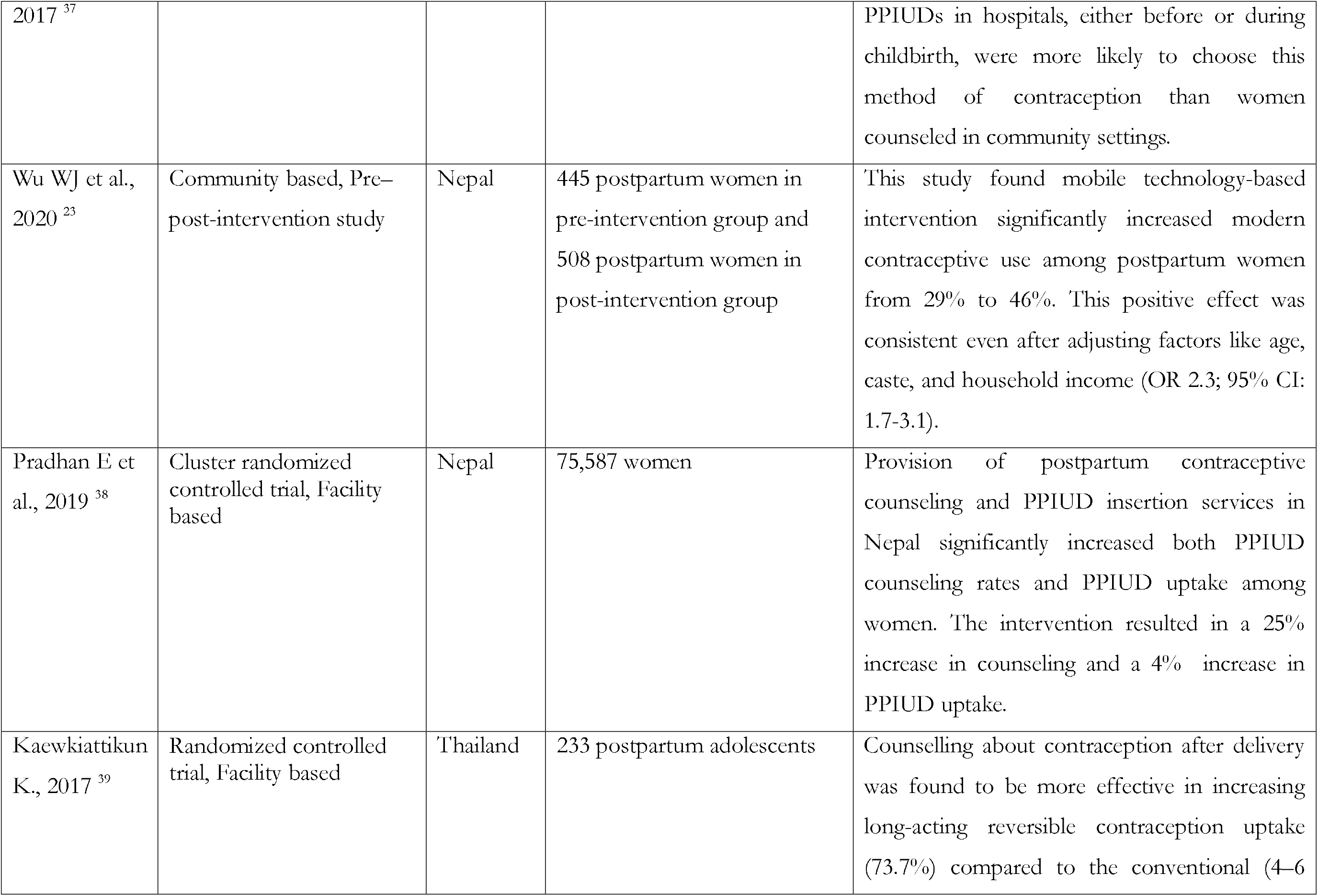
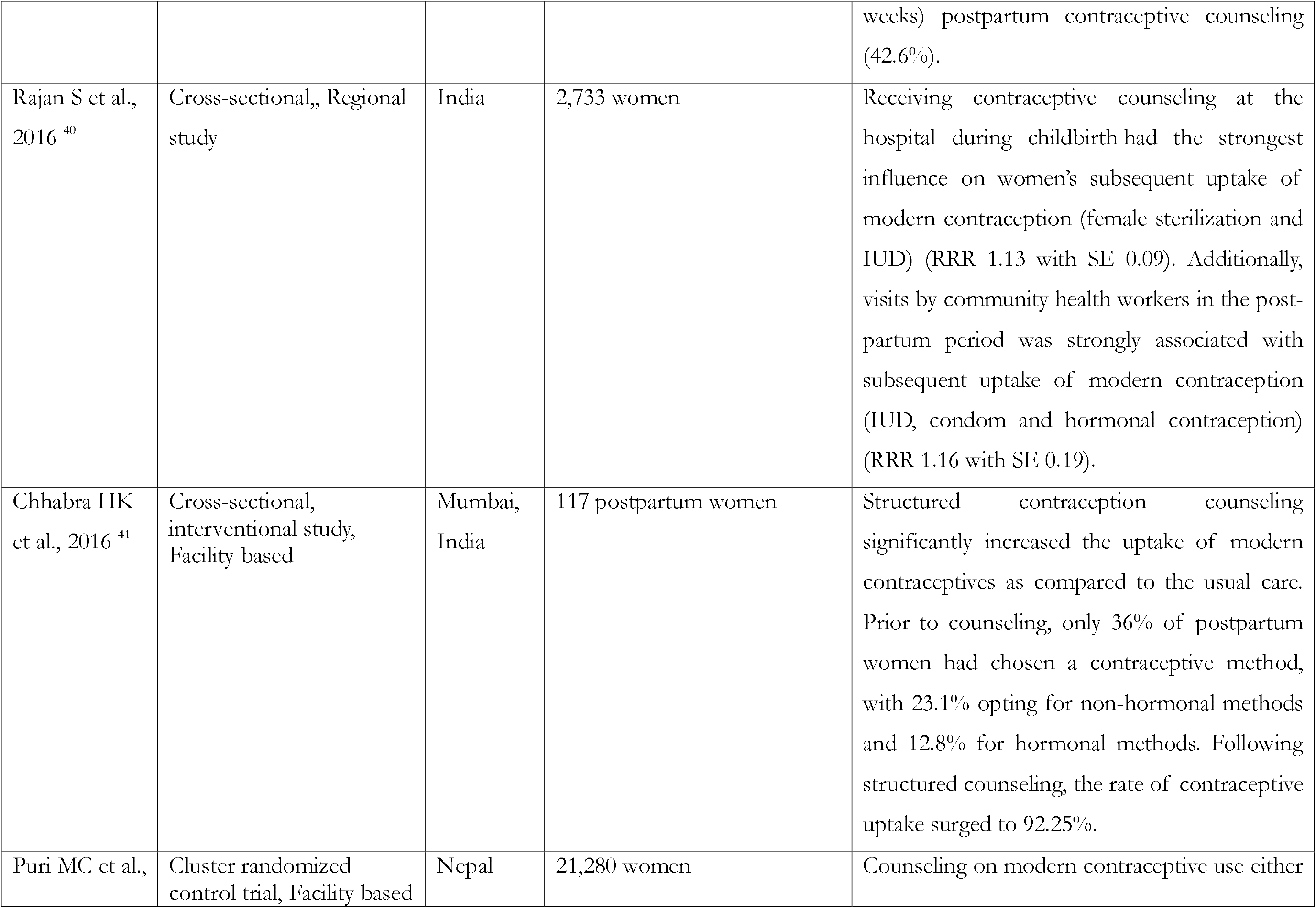
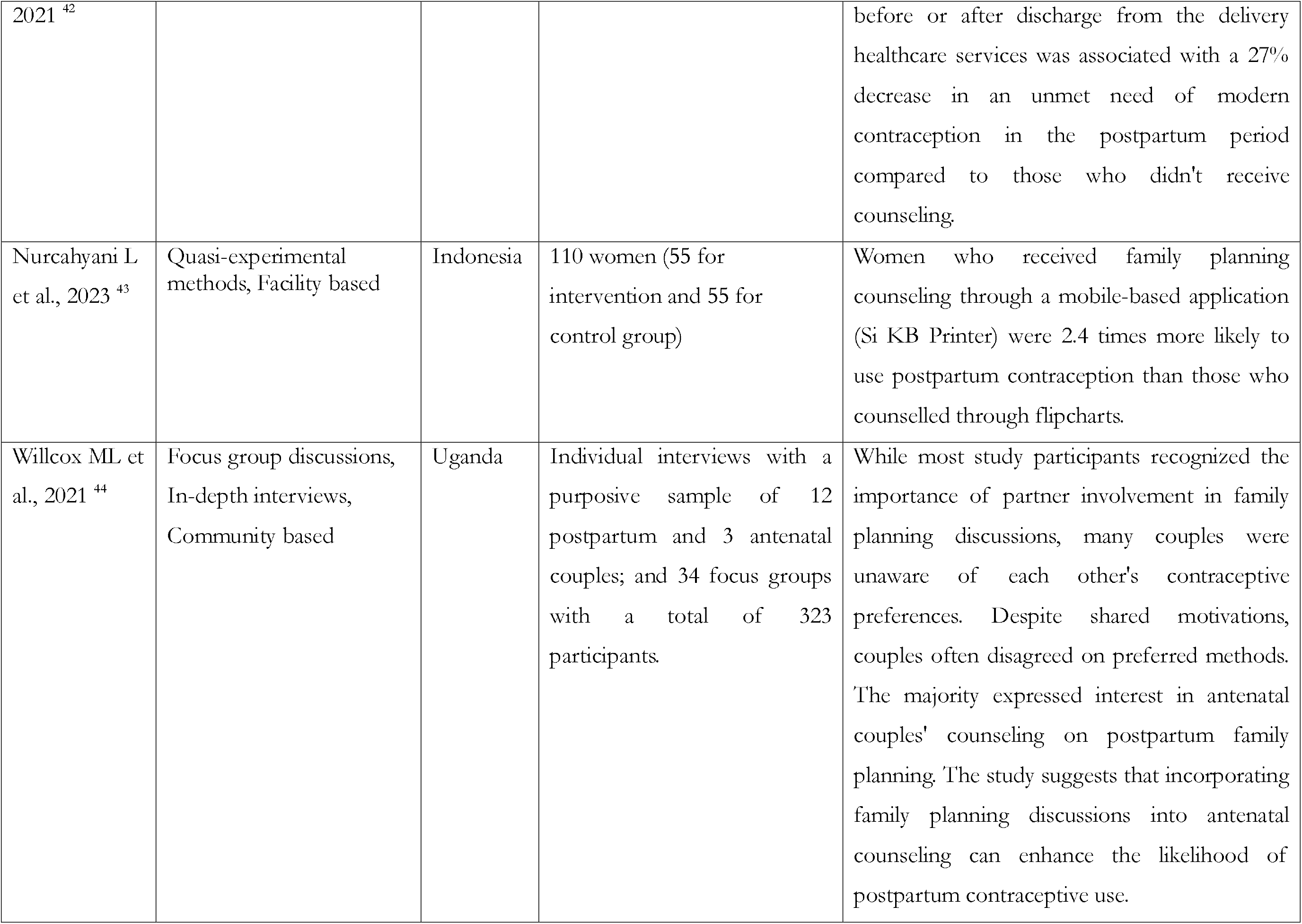
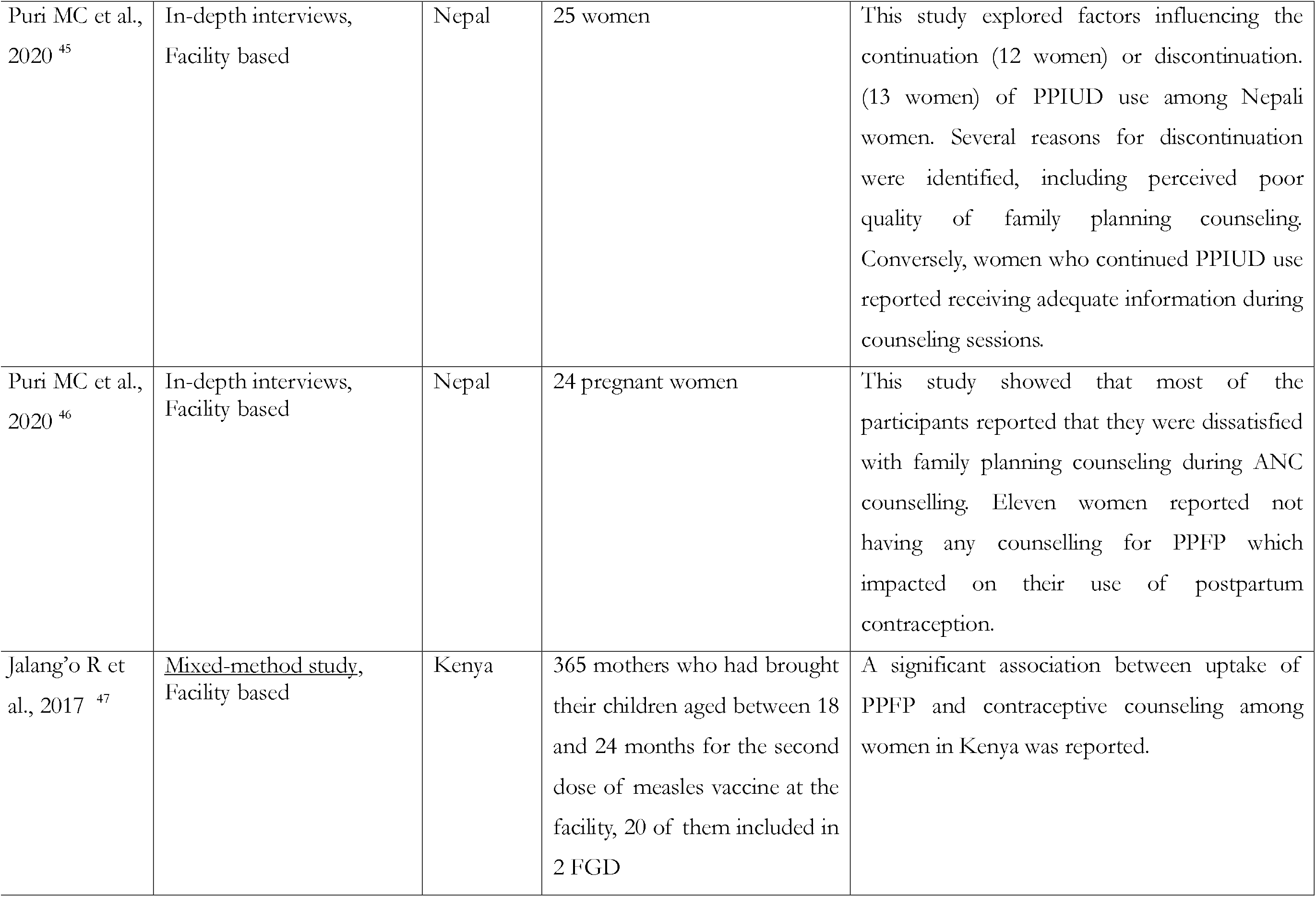
Narrative review of family planning counselling and postpartum contraceptive uptake for the articles that did not follow the WHO classification in low- and lower-middle-income countries, January 2015 to April 2024

#### Narrative synthesis of studies assessing the effects of family planning counseling on modern contraception uptake during the postpartum period

Our narrative synthesis included 19 studies, which produced results consistent with the main summary estimates. All of these studies provided evidence of a significant increase in modern contraception uptake during the postpartum period following counseling received during maternal healthcare services.

## Discussion

This study explored the effects of family planning counselling during any point of engagement with maternal healthcare on the uptake of modern contraception in the postpartum period. Based on a summary estimate from 42 studies, we found that women who being counselled on family planning services during maternal healthcare services uptake were 2.75 times (95% CI, 2.11-3.58) more likely to use modern contraception during the postpartum period compared to those who did not receive counseling. Although we observed significant publication bias and heterogeneity, addressing these issues through trim and fill estimation and subgroup analyses, respectively, produced estimates consistent with the summary findings. A narrative synthesis of an additional 19 studies also supports the effectiveness of family planning counseling during maternal healthcare on postpartum contraception uptake. Together, these findings provide a strong basis for concluding that family planning counseling during maternal healthcare effectively increases the uptake of modern contraception during the postpartum period. This evidence suggests the need for policies and programs to scale up family planning counseling as part of maternal healthcare services.

The observed increase in postpartum contraceptive use among women who receive family planning counselling can be attributed to several key factors ^25,48^. Family planning counselling provides comprehensive information on various contraceptive options, including their effectiveness, side effects, and usage instructions ^21^. This education empowers women to make informed reproductive health decisions, which facilitates the correct use of modern contraception during the postpartum period ^49,50^. Moreover, family planning counselling also raises awareness about the benefits of postpartum contraception, such as improved birth spacing and the prevention of unintended pregnancies ^17,51^. This increased awareness promotes the role of contraception in enhancing maternal and child health, thereby encouraging its uptake ^21,51^. Additionally, family planning counselling sessions offer an opportunity to dispel myths and address misconceptions about contraception (e.g, breastfeeding work as effective contraception), reducing anxiety and promoting its adoption ^52^.

Personalized family planning counselling is another critical element, allowing healthcare providers to consider a woman’s individual health status, reproductive goals, and personal circumstances ^52-54^. Tailored advice leads to higher satisfaction and continuation rates. The ongoing nature of family planning counselling during maternal healthcare services uptake builds trust and rapport between women and their healthcare providers, making women more comfortable discussing contraception and more likely to follow medical advice ^53,55^. Moreover, repeated family planning counselling during prenatal and postnatal visits reinforces key health messages, increasing the likelihood of contraceptive adoption and continuation ^21,56^. Integrating family planning counseling into maternal healthcare reduces barriers such as the need for additional appointments or travel, making contraceptive access more convenient ^57^. Furthermore, family planning counselling sessions often involve partners in family planning discussions, which can significantly increase support for contraceptive use ^19^. Normalizing contraception as a standard part of postpartum care further encourages its adoption ^48,57^. Family planning counselling also helps identify women with unmet contraceptive needs, addressing these during routine maternal healthcare visits ^42^.

Although our additional analyses yielded estimates consistent with the summary findings, they provide critical insights. Evidence of publication bias addressed through the trim and fill method indicates 12 missing studies with a reverse association between family planning counseling and modern contraception uptake during the postpartum period. This may be due to a tendency among authors and journals to publish studies with positive associations. Subgroup analyses across confounding factors indicate no association between family planning counseling and modern contraception uptake during the postpartum period, which becomes significantly positive once confounders are adjusted. This underscores the role of socio-demographic factors in shaping the observed association. Particularly, these factors influence whether women access maternal healthcare services, despite governmental efforts in LMICs to achieve universal healthcare coverage as part of the SDGs. Women who do not access maternal healthcare services represent missed opportunities for healthcare providers to counsel on modern contraception during the postpartum period ^58^. We also found that family planning counselling at each stage of maternal healthcare effectively promotes modern contraception uptake during the postpartum period ^48,59-66^. However, family planning counselling received during delivery healthcare was found to be more effective, though this finding is based on a single study ^67^. The underlying reason is that postpartum contraception issues arise immediately following delivery healthcare rather than antenatal healthcare. Moreover, some LMICs, such as Bangladesh, have policies to supply long-acting contraception to women during delivery healthcare services ^13,68,69^. Family planning counselling during postnatal care can play an even more effective role ^62,63,70,71^. However, in LMICs, mothers who deliver in healthcare facilities usually receive their first postnatal healthcare before discharge, with subsequent postnatal healthcare services often considered less important unless complications arise ^72,73^. Moreover, the insignificant association reported for countries, such as Uganda, may be due to lower utilization of maternal healthcare services and the lack of options for family planning counseling, or a combination of both ^74,75^. However, further exploration is necessary for these countries.

The findings of this study have significant policy implications, highlighting the need to integrate family planning counseling as a standard component of maternal healthcare services. This can be achieved by training healthcare providers, equipping them with the necessary skills and knowledge to deliver effective family planning counseling, and developing and implementing standardized protocols to ensure consistent and high-quality information delivery. Additionally, identifying and addressing potential barriers that may prevent women from accessing or utilizing counseling services is crucial. This might involve considering cultural sensitivities, language accessibility, and childcare options during appointments.

This study has several key strengths and some limitations. To our knowledge, this is the first study in LMICs that summarizes the effects of family planning counseling during maternal healthcare services on modern contraception uptake during the postpartum period. We included all studies conducted in LMICs since 2015, when the SDGs were established to reduce maternal and child mortality, in either quantitative or narrative synthesis. We used comprehensive search techniques for data extraction and selection of eligible studies and followed the “Strengthening the Reporting of Observational Studies in Epidemiology” (STROBE) and PRISMA guidelines for reporting study findings. Additionally, we considered family planning counseling received at any stage of maternal healthcare services and analyzed them both combined and separately. A summary estimate was calculated using random effects meta-analysis, and heterogeneity was addressed through subgroup analyses. However, the summary estimates presented in this study were mainly based on cross-sectional studies. We did not search for unpublished papers and grey literature, which could contribute to publication bias. However, this bias was addressed through the trim and fill method, and similar results were found, suggesting that our findings are reliable. We considered family planning counseling in general, although it may take several forms, such as direct discussion or providing handouts along with discussions during maternal healthcare services. Unfortunately, we could not differentiate the types of counseling received due to a lack of relevant data in the included studies. Moreover, we used the broad term “modern contraception method” for all types of modern contraception, regardless of their varying effectiveness. However, a lack of data restricted us from making such distinctions.

## Conclusion

This study offers robust evidence that integrating family planning counseling into maternal healthcare significantly increases postpartum contraceptive use. This finding underscores the critical role of comprehensive counseling throughout the entire spectrum of maternal care. By empowering women with knowledge and options, we can enable them to make informed choices about their reproductive health and well-being.

## Supporting information

Linked

## Data Availability

All data produced in the present work are contained in the manuscript

## Acknowledgements

We acknowledge the support of the Department of Population Science of the Jatiya Kabi Kazi Nazrul Islam University, Bangaladesh where this research was conducted.

## Authorship contributions

MNK and MLH developed the study concept. ARC and MSR reviewed the articles independently, extracted data, and assessed study quality. MNK conducted the formal analysis drafted the manuscript. ARC and RH performed the quality assessment and checked the results. TAH, CC and MLH critically reviewed and edited all previous versions of the manuscript. All authors approved the final version of this manuscript.

