## Supplementary material for "Effects of family planning counseling delivered during maternal healthcare on postpartum modern contraceptive uptake in low- and middle-income countries: a systematic review and meta-analysis": Linked

**Supplementary Table 01.** Search strategy of Medline database (date: January 2015 to April 2024)

| **#** | **Searches** | **Results** |
| --- | --- | --- |
| 1 | (Family planning counseling OR family planning information OR family planning advice OR contraceptive counseling).mp | 1,617 |
| 2 | (Postpartum* OR post-partum* OR Postnatal*).ti,ab. OR (Postpartum* OR post-partum* OR Postnatal*).sh. | 188,604 |
| 3 | (contraceptive* OR family planning OR birth control OR depo? Medroxyprogesterone OR Depo-Provera OR Sayana Press OR IUD OR IUCD OR IUS OR intrauterine device* OR intrauterine system* OR oral contraceptive pill* OR hormonal contraceptive pill* OR birth control pill OR emergency contraceptive pill*OR cervical cap* OR vaginal diaphragm* OR vaginal ring* OR implant* OR subdermal implant* OR implanon OR jadelle OR norplant* OR sinoimplant OR sterilization OR vasectomy OR contraception behavior OR long-acting reversible contraception).ti,ab. OR family planning/ OR birth control/ OR contraception/ OR Contraception Behavior/ OR Family Planning Services/ | 532,994 |
| 4 | (Use* OR uptake OR utilization* OR adoption* OR initiation*).ti,ab. OR (Use* OR uptake OR utilization* OR adoption* OR initiation*).sh. | 7,889,916 |
| 5 | (Angola* OR Benin* OR Botswana* OR Burkina Faso* OR Burundi* OR Cabo Verde* OR Cameroon* OR Central African Republic* OR Chad* OR Comoros* OR Congo* OR Cote D'ivoire* OR Ivory coast* OR Guinea* OR Eritrea* OR Eswatini* OR Swaziland* OR Ethiopia* OR Gabon* OR Gambia* OR Ghana* OR Guinea* OR Guinea-Bissau* OR Kenya* OR Lesotho* OR Liberia* OR Madagascar* OR Malawi* OR Mali* OR Mauritania* OR Mauritius* OR Mozambique* OR Namibia* OR Niger* OR Nigeria* OR Rwanda* OR (Sao Tome and Principe*) OR Senegal* OR Seychelles* OR Sierra Leone* OR Somalia* OR South Africa* OR South Sudan* OR Sudan* OR Tanzania* OR Togo* OR Uganda* OR Zambia* OR Zimbabwe*).ti,ab. OR (sub-Saharan Africa OR Africa OR East Africa OR West Africa OR South Africa OR Central Africa).ti,ab. OR (sub-Saharan Africa OR Africa OR East Africa OR West Africa OR South Africa OR Central Africa).sh. | 1,053,931 |
| 6 | 2 and 3 and 4 | 3,268 |
| 7 | 1 or 6 | 4,698 |
| 8 | 7 and 5 | 841 |
| 9 | (HIV or adolescent or postabortion or post-abortion or refugee or migrant).mp. | 2,443,191 |
| 10 | 8 not 9 | 394 |
| 11 | Limit 10 to English language | 370 |

**Supplementary Table 02.** Search strategy of Embase database (date: January 2015 to April 2024)

| **#** | **Searches** | **Results** |
| --- | --- | --- |
| 1 | (Family planning counseling OR family planning information OR family planning advice OR contraceptive counseling).mp. | 1951 |
| 2 | (Postpartum* OR post-partum* OR Postnatal*).ti,ab. OR (Postpartum* OR post-partum* OR Postnatal*).sh. | 237,172 |
| 3 | (contracepti* OR family planning OR birth control OR depo? Medroxyprogesterone OR Depo-Provera OR Sayana Press OR IUD OR IUCD OR IUS OR intrauterine device* OR intrauterine system* OR oral contraceptive pill* OR hormonal contraceptive pill* OR birth control pill OR emergency contraceptive pill*OR cervical cap* OR vaginal diaphragm* OR vaginal ring* OR implant* OR subdermal implant* OR implanon OR jadelle OR norplant* OR sinoimplant OR sterilization OR vasectomy OR contraception behavior OR long-acting reversible contraception).ti,ab. OR family planning/ OR birth control/ OR contraception/ OR Contraception Behavio?r/ OR Family Planning Services/ | 713,261 |
| 4 | (Use* OR uptake OR utilization* OR adoption* OR initiation*).ti,ab. OR (Use* OR uptake OR utilization* OR adoption* OR initiation*).sh. | 10,262,706 |
| 5 | (Angola* OR Benin* OR Botswana* OR Burkina Faso* OR Burundi* OR Cabo Verde* OR Cameroon* OR Central African Republic* OR Chad* OR Comoros* OR Congo* OR Cote D'ivoire* OR Ivory coast* OR Guinea* OR Eritrea* OR Eswatini* OR Swaziland* OR Ethiopia* OR Gabon* OR Gambia* OR Ghana* OR Guinea* OR Guinea-Bissau* OR Kenya* OR Lesotho* OR Liberia* OR Madagascar* OR Malawi* OR Mali* OR Mauritania* OR Mauritius* OR Mozambique* OR Namibia* OR Niger* OR Nigeria* OR Rwanda* OR (Sao Tome and Principe*) OR Senegal* OR Seychelles* OR Sierra Leone* OR Somalia* OR South Africa* OR South Sudan* OR Sudan* OR Tanzania* OR Togo* OR Uganda* OR Zambia* OR Zimbabwe*).ti,ab. OR (sub-Saharan Africa OR Africa OR East Africa OR West Africa OR South Africa OR Central Africa).ti,ab. OR (sub-Saharan Africa OR Africa OR East Africa OR West Africa OR South Africa OR Central Africa).sh. | 1,351,451 |
| 6 | 2 and 3 and 4 | 4,004 |
| 7 | 1 or 6 | 5,700 |
| 8 | 7 and 5 | 859 |
| 9 | (HIV or adolescent or postabortion or post-abortion or refugee or migrant).mp. | 2,036,452 |
| 10 | 8 not 9 | 420 |
| 11 | Limit 10 to English language | 409 |

**Supplementary Table 03.** Search strategy of Global Health database (date: January 2015 to April 2024)

| **#** | **Searches** | **Results** |
| --- | --- | --- |
| 1 | (Family planning counseling OR family planning information OR family planning advice OR contraceptive counseling).mp. | 424 |
| 2 | (Postpartum* OR post-partum* OR Postnatal*).ti,ab. OR (Postpartum* OR post-partum* OR Postnatal*).sh. | 27,598 |
| 3 | (contraception* OR family planning OR birth control OR depo? Medroxyprogesterone OR Depo-Provera OR Sayana Press OR IUD OR IUCD OR IUS OR intrauterine device* OR intrauterine system* OR oral contraceptive pill* OR hormonal contraceptive pill* OR birth control pill OR emergency contraceptive pill*OR cervical cap* OR vaginal diaphragm* OR vaginal ring* OR implant* OR subdermal implant* OR implanon OR jadelle OR norplant* OR sinoimplant OR sterilization OR vasectomy OR contraception behavior OR long-acting reversible contraception).ti,ab. OR family planning/ OR birth control/ OR contraception/ OR Contraception Behavior/ OR Family Planning Services/ | 35,767 |
| 4 | (Use* OR uptake OR utilization* OR adoption* OR initiation*).ti,ab. OR (Use* OR uptake OR utilization* OR adoption* OR initiation*).sh. | 1,286,816 |
| 5 | (Angola* OR Benin* OR Botswana* OR Burkina Faso* OR Burundi* OR Cabo Verde* OR Cameroon* OR Central African Republic* OR Chad* OR Comoros* OR Congo* OR Cote D'ivoire* OR Ivory coast* OR Guinea* OR Eritrea* OR Eswatini* OR Swaziland* OR Ethiopia* OR Gabon* OR Gambia* OR Ghana* OR Guinea* OR Guinea-Bissau* OR Kenya* OR Lesotho* OR Liberia* OR Madagascar* OR Malawi* OR Mali* OR Mauritania* OR Mauritius* OR Mozambique* OR Namibia* OR Niger* OR Nigeria* OR Rwanda* OR (Sao Tome and Principe*) OR Senegal* OR Seychelles* OR Sierra Leone* OR Somalia* OR South Africa* OR South Sudan* OR Sudan* OR Tanzania* OR Togo* OR Uganda* OR Zambia* OR Zimbabwe*).ti,ab. OR (sub-Saharan Africa OR Africa OR East Africa OR West Africa OR South Africa OR Central Africa).ti,ab. OR (sub-Saharan Africa OR Africa OR East Africa OR West Africa OR South Africa OR Central Africa).sh. | 302,980 |
| 6 | 2 and 3 and 4 | 883 |
| 7 | 1 or 6 | 1,249 |
| 8 | 7 and 5 | 505 |
| 9 | (HIV or adolescent or postabortion or post-abortion or refugee or migrant).mp. | 215,725 |
| 10 | 8 not 9 | 320 |
| 11 | Limit 10 to English language | 323 |

**Supplementary Table 04.** Search strategy of Scopus database (date: January 2015 to April 2024)

| **#** | **Searches** | **Results** |
| --- | --- | --- |
| #1 | TITLE-ABS-KEY ("Family planning counseling*" OR "family planning information" OR "family planning advice" OR  "contraceptive counseling*") | 760 |
| #2 | TITLE-ABS-KEY (“Postpartum*” OR “post-partum*” OR “postnatal*”) | 226,414 |
| #3 | TITLE-ABS-KEY (contraceptive* OR "family planning" OR "birth control" OR "depo? Medroxyprogesterone" OR "Depo-Provera "OR "Sayana Press" OR "IUD" OR "IUCD*" OR "IUS" OR "intrauterine device*" OR "intrauterine system*" OR "ral contraceptive pill*" OR "hormonal contraceptive pill*" OR "birth control pill" OR "emergency contraceptive pill*" OR "cervical cap*" OR "vaginal diaphragm*" OR "vaginal ring*" OR "implant*" OR "subdermal implant*" OR "implanon" OR "jadelle" OR "norplant*" OR "sinoimplant*" OR "sterilization" OR "vasectomy" OR "contraception behavior" OR "long-acting reversible contraception") | 1,041,555 |
| #4 | TITLE-ABS-KEY ("Use*" OR "uptake" OR "utilization*" OR "adoption*" OR "initiation*") | 22,782,654 |
| # | TITLE-ABS-KEY (Angola* OR Benin* OR Botswana* OR "Burkina Faso*" OR Burundi* OR "Cabo Verde*" OR Cameroon* OR "Central African Republic*" OR Chad* OR Comoros* OR Congo* OR "Cote D'ivoire*" OR "Ivory coast*" OR Guinea* OR Eritrea* OR Eswatini* OR Swaziland* OR Ethiopia* OR Gabon* OR Gambia* OR Ghana* OR Guinea* OR "Guinea-Bissau*" OR Kenya* OR Lesotho* OR Liberia* OR Madagascar* OR Malawi* OR Mali* OR Mauritania* OR Mauritius* OR Mozambique* OR Namibia* OR Niger* OR Nigeria* OR Rwanda* OR "Sao Tome and Principe*" OR Senegal* OR Seychelles* OR "Sierra Leone*" OR Somalia* OR "South Africa*" OR "South Sudan*" OR Sudan* OR Tanzania* OR Togo* OR Uganda* OR Zambia* OR Zimbabwe* OR "sub-Saharan Africa" OR Africa OR "East Africa" OR "West Africa" OR "South Africa" OR "Central Africa") | 2,020,233 |
| #6 | #2 AND #3 AND #4 | 4,407 |
| #7 | #1 OR #6 | 5,093 |
| #8 | #7 AND #5 | 863 |
| #9 | #8 AND NOT (HIV OR adolescent OR postabortion OR post-abortion OR refugee OR migrant) AND (LIMIT-TO (LANGUAGE," English")) | 290 |
| #10 | #9 AND (LIMIT-TO ( AFFILCOUNTRY ,  "Nigeria" )  OR  LIMIT-TO ( AFFILCOUNTRY ,  "Ethiopia" )  OR  LIMIT-TO ( AFFILCOUNTRY ,  "South Africa" )  OR  LIMIT-TO ( AFFILCOUNTRY ,  "Kenya" )  OR  LIMIT-TO ( AFFILCOUNTRY ,  "Tanzania" )  OR  LIMIT-TO ( AFFILCOUNTRY ,  "Uganda" )  OR  LIMIT-TO ( AFFILCOUNTRY ,  "Ghana" )  OR  LIMIT-TO ( AFFILCOUNTRY ,  "Malawi" )  OR  LIMIT-TO ( AFFILCOUNTRY ,  "Burkina Faso" )  OR  LIMIT-TO ( AFFILCOUNTRY ,  "Rwanda" )  OR  LIMIT-TO ( AFFILCOUNTRY ,  "Zambia" )  OR  LIMIT-TO ( AFFILCOUNTRY ,  "Congo" )  OR  LIMIT-TO ( AFFILCOUNTRY ,  "Democratic Republic Congo" )  OR  LIMIT-TO ( AFFILCOUNTRY ,  "Guinea" )  OR  LIMIT-TO ( AFFILCOUNTRY ,  "Sudan" )  OR  LIMIT-TO ( AFFILCOUNTRY ,  "Zimbabwe" )  OR  LIMIT-TO ( AFFILCOUNTRY ,  "Botswana" )  OR  LIMIT-TO ( AFFILCOUNTRY ,  "Cameroon" )  OR  LIMIT-TO ( AFFILCOUNTRY ,  "Liberia" )  OR  LIMIT-TO ( AFFILCOUNTRY ,  "Mali" )  OR  LIMIT-TO ( AFFILCOUNTRY ,  "Mozambique" )  OR  LIMIT-TO ( AFFILCOUNTRY ,  "Senegal" )  OR  LIMIT-TO ( AFFILCOUNTRY ,  "Sierra Leone" )  OR  LIMIT-TO ( AFFILCOUNTRY ,  "Undefined" ) ) | 217 |

**Supplementary Table 05.** Search strategy of PubMed database (date: January 2015 to April 2024)

| **#** | **Searches** | **Results** |
| --- | --- | --- |
| #1 | Family planning counselling[tiab] OR family planning counselling[mh] OR family planning counseling[tiab] OR family planning counseling[mh] OR family planning information [tiab] OR family planning advice[tiab] contraceptive counselling[tiab] OR contraceptive counselling[mh] OR contraceptive counseling[tiab] OR contraceptive counseling[mh] | 2,950 |
| #2 | Postpartum*[tiab] OR Postpartum*[mh] OR post-partum*[tiab] OR post-partum*[mh] or postnatal*[tiab] or postnatal*[mh] | 231,117 |
| #3 | contraceptive*[tiab] OR family planning[tiab] OR family planning[mh] OR birth control[tiab] OR birth control[mh] OR Depot Medroxyprogesterone[tiab] OR depomedroxyprogesterone[tiab] OR Depo-Provera[tiab] OR Sayana Press[tiab] OR IUD[tiab] OR IUCD[tiab] OR IUS OR intrauterine device*[tiab] OR intra-uterine device*[tiab] OR intrauterine system*[tiab] OR intra-uterine system*[tiab] OR oral contraceptive pill*[tiab] OR hormonal contraceptive pill*[tiab] OR birth control pill[tiab] OR emergency contraceptive pill*[tiab] OR cervical cap*[tiab] OR vaginal diaphragm*[tiab] OR vaginal ring*[tiab] OR implant*[tiab] OR subdermal implant*[tiab] OR implanon[tiab] OR jadelle[tiab] OR norplant*[tiab] OR sino-implant*[tiab] OR sinoimplant*[tiab] OR sterilization[tiab] OR sterilisation[tiab] OR vasectomy[tiab] OR contraception behaviour[tiab] OR long-acting reversible contraception[tiab] OR family planning[mh] OR birth control[mh] OR contraception[mh] OR Family Planning Services[mh] | 544,236 |
| #4 | Use[tiab] OR uptake[tiab] OR utilization[tiab] OR utilisation[tiab] OR adoption[tiab] OR initiation[tiab] OR Use[mh] OR uptake[mh] OR utilization[mh] OR utilisation[mh] OR adoption[mh] OR initiation[mh] | 3,677,906 |
| #5 | Angola*[tiab] OR Benin* [tiab] OR Botswana*[tiab] OR Burkina Faso*[tiab] OR Burundi*[tiab] OR Cabo Verde*[tiab] OR Cameroon*[tiab] OR Central African Republic*[tiab] OR Chad*[tiab] OR Comoros*[tiab] OR Congo*[tiab] OR Cote D'ivoire*[tiab] OR Ivory coast*[tiab] OR Guinea*[tiab] OR Eritrea*[tiab] OR Eswatini*[tiab] OR Swaziland*[tiab] OR Ethiopia*[tiab] OR Gabon*[tiab] OR Gambia*[tiab] OR Ghana*[tiab] OR Guinea*[tiab] OR Guinea-Bissau*[tiab] OR Kenya*[tiab] OR Lesotho*[tiab] OR Liberia*[tiab] OR Madagascar*[tiab] OR Malawi*[tiab] OR Mali*[tiab] OR Mauritania*[tiab] OR Mauritius*[tiab] OR Mozambique*[tiab] OR Namibia*[tiab] OR Niger*[tiab] OR Nigeria*[tiab] OR Rwanda*[tiab] OR Sao Tome and Principe*[tiab] OR Senegal*[tiab] OR Seychelles*[tiab] OR Sierra Leone*[tiab] OR Somalia*[tiab] OR South Africa*[tiab] OR South Sudan*[tiab] OR Sudan*[tiab] OR Tanzania*[tiab] OR Togo*[tiab] OR Uganda*[tiab] OR Zambia*[tiab] OR Zimbabwe*[tiab] OR sub-Saharan Africa[tiab] OR sub-Saharan Africa[mh] OR Africa[tiab] OR Africa[mh] OR East Africa[tiab] OR East Africa[mh] OR West Africa[tiab] OR West Africa[mh] OR South Africa[tiab] OR South Africa[mh] OR Central Africa[tiab] OR Central Africa[mh] | 1,121,789 |
| #6 | HIV OR Adolescent OR postabortion OR post-abortion OR migrant OR refugee | 2,715,880 |
| #7 | #2 AND #3 AND #4 | 2,789 |
| #8 | #1 OR #7 | 5,545 |
| #9 | #8 AND 5 | 958 |
| #10 | #7 AND #5 Filters:English | 924 |
| #11 | #10 NOT #6 Filters:English | 345 |

**Supplementary Table 06.** Search strategy of Web Science database (date: January 2015 to April 2024)

| **#** | **Searches** | **Results** |
| --- | --- | --- |
| #1 | TS= ("Family planning counseling*" OR "family planning information" OR "family planning advice" OR "contraceptive counseling*") | 395 |
| #2 | TS= ("Postpartum*" OR "post-partum*" OR "postnatal*") | 183,159 |
| #3 | TS= (contraceptive* OR "family planning" OR "birth control" OR "depo? Medroxyprogesterone" OR "Depo-Provera "OR "Sayana Press" OR "IUD" OR "IUCD*" OR "IUS" OR "intrauterine device*" OR "intrauterine system*" OR "oral contraceptive pill*" OR "hormonal contraceptive pill*" OR "birth control pill" OR "emergency contraceptive pill*" OR "cervical cap*" OR "vaginal diaphragm*" OR "vaginal ring*" OR "implant*" OR "subdermal implant*" OR "implanon" OR "jadelle" OR "norplant*" OR "sinoimplant*" OR "sterilization" OR "vasectomy" OR "contraception behavior" OR "long-acting reversible contraception") | 638,837 |
| #4 | TS= (“Use*” OR “uptake” OR “utilization*” OR “adoption*” OR “initiation*”) | 15,774,374 |
| #5 | TS= (Angola* OR Benin* OR Botswana* OR "Burkina Faso*" OR Burundi* OR "Cabo Verde*" OR Cameroon* OR "Central African Republic*" OR Chad* OR Comoros* OR Congo* OR "Cote D'ivoire*" OR "Ivory coast*" OR Guinea* OR Eritrea* OR Eswatini* OR Swaziland* OR Ethiopia* OR Gabon* OR Gambia* OR Ghana* OR Guinea* OR "Guinea-Bissau*" OR Kenya* OR Lesotho* OR Liberia* OR Madagascar* OR Malawi* OR Mali* OR Mauritania* OR Mauritius* OR Mozambique* OR Namibia* OR Niger* OR Nigeria* OR Rwanda* OR "Sao Tome and Principe*" OR Senegal* OR Seychelles* OR "Sierra Leone*" OR Somalia* OR "South Africa*" OR "South Sudan*" OR Sudan* OR Tanzania* OR Togo* OR Uganda* OR Zambia* OR Zimbabwe* OR "sub-Saharan Africa" OR Africa OR "East Africa" OR "West Africa" OR "South Africa" OR "Central Africa") | 1,537,528 |
| #7 | #2 AND #3 AND #4 | 2,567 |
| #8 | #1 OR #7 | 2,917 |
| #9 | #8 AND #5 | 530 |
| #10 | #9 AND English | 526 |
| #11 | TS=(HIV OR adolescent OR postabortion OR "post-abortion" OR refugee OR migrant) | 872,020 |
| #12 | #10 NOT #11 | 350 |

**Supplementary Table 07**. Summary of selected observational studies on family planning counselling and postpartum modern contraceptive uptake in low- and lower-middle income countries, January 2015 to April, 2024.

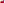

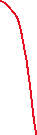

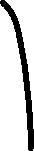

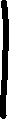

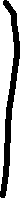

| **Authors, year, and country** | **Study design and settings** | **Sample** | **Outcomes** | **Confounders adjustment** |
| --- | --- | --- | --- | --- |
| Arero WD et al., 2022, Ethiopia ^1^ | Cross sectional, Facility-based study | 393 | Long-acting reversible contraceptive use | Maternal age, mode of delivery, current birth outcome, number of parity, number of alive kids, completed family size, next delivery plan within two years, ever heard of a LARC, Previous LARC use, monthly income |
| Hagos KN et al.,2023  Ethiopia ^2^ | Case control, Facility-based study | 308 | Postpartum contraceptive use | Maternal age, residence, ANC visits, still birth |
| Murry LL et al.,2021, India ^3^ | Cross sectional, Facility-based study | 284 | Contraceptive use | Religion, education, number of ANC visit, breast feeding status, resume menses after birth, resumed sexual activity |
| Mruts KB et al.,2023, Ethiopia ^4^ | Prospective cohort, National study | 769 | Unmet need for contraception | Child immunization |
| Adofo E et al.,2021, Ghana ^5^ | Cross sectional, Facility-based study | 422 | Contraceptive use | Age, education, occupation, resumption of menses, residence, age of child, fear of infertility, agreed to contraceptive use on ANC counselling, Partners discussion on contraceptive |
| Wudineh KG et al.,2023, Ethiopia ^6^ | Cross sectional , Facility-based study | 417 | Immediate postpartum reversible long-acting contraceptive uptake | Age, residency, discussion with partner, ANC follow-up, attitude to FP, outcome of neonate, plan to conceive next pregnancy, history of FP use |
| Melkie A et al.,2021, Ethiopia ^7^ | Cross sectional, Facility-based study | 423 | Utilization of IPPIUCD | Age, education, birth spacing |
| Kiondo SK et al.,2020, Tanzania ^8^ | Cross sectional, Community based study | 141 | LARC use | Age, marital status, residence, occupation, income per month and Partner’s level of education, Number of living children, mode of index delivery, place of index delivery, menstrual resumption, duration of postpartum period and discussing FP use with partner. |
| Silesh M et al.,2022, Ethiopia ^9^ | Cross sectional, Facility-based study | 394 | IPPFP utilisation | age, residence, women’s educational status, husband’s educational status, ever used FP, partner support to use FP, knowledge on FP attitude towards FP, history of abortion, mode of delivery, current birth outcome, planning status of history of ANC for current pregnancy, and maternal satisfaction with intrapartum care. |
| Nugussa B et al.,2023, Ethiopia ^10^ | Cross sectional , Community based study | 385 | Utilization of modern postpartum family planning | Age of current child, planned birth, parity, condition of menses, visiting HF  after delivery, ever used modern FP methods, Decision of FP use, husband approval on FP use, discussion with a partner about FP |
| Maeregayehu T et al.,2022, Ethiopia ^11^ | Retrospective cohort, Community based study | 594 | Use of postpartum modern contraceptive | educational status of women, occupation status of women, Number of ANC visit, Distance to the health facility, FP counseling during child immunization, fertility desire, resume sexual intercourse, resume menses, discuss with the partner on FP, who decide on PPFP use, Knowledge of Postpartum family planning. |
| Tilahun T et al.,2022, Ethiopia ^12^ | Cross sectional, Facility-based study | 989 | Postpartum family planning uptake | Parity, FP utilization before the index pregnancy, previous history of ppfp uptake, place of delivery, skilled birth attendance, Faced difficulty during COVID-19 restrictions, ANC during the index pregnancy |
| Getaneh M et al.,2021, Ethiopia ^13^ | Cross sectional, Community based study | 630 | Extended postpartum period modern contraceptive utilization | educational status, discussion with husband, link to family planning during child immunization, sexual resumption, menstrual resumption, fertility desire |
| Agula C et al.,2022  , Ghana ^14^ | Cross sectional, Regional study | 624 | Modern contraceptive use in postpartum period | Age, marital status, education, religion, number of live births, Attended postnatal care after birth |
| Abebe GF et al.,2023  , Ethiopia ^15^ | Cross sectional, Community based study | 385 | Utilization of modern contraceptives | Education, husband’s education, residence, number of children, heard about FP, ANC follow-up, contraceptive used previously, started menstruation, resumed sexual intercourse |
| Ezekiel MJ et al.,2022  , Tanzania ^16^ | Cross sectional, Regional study | 299 | Taking FP method | Occupation, mode of payment, parity, number of ANC visit |
| Tafa L et al.,2021  , Ethiopia ^17^ | Cross sectional, Facility-based study | 625 | PPFP use | Previous FP information, FP information from health facility visit, ANC attendance, menses resumption, Sexual resumption |
| Nakiwunga N et al.,2022  , Uganda ^18^ | Cross sectional, Facility-based study | 397 | Contraceptive uptake | Parity, ever had abortion, children alive, Mode of delivery, birth interval, planned time for next pregnancy, Number of ANC, ever used contraception, Method used before |
| Abraha TH et al.,2017  , Ethiopia ^19^ | Cross sectional, Community based study | 590 | Postpartum modern contraceptive use | Education, parity, living children, currently breastfeed, ANC, PNC, Postpartum period (wk), Discussed FP with husband, husband approval of FP, Knowledge of postpartum FP, Received FP counseling by health extension work, experienced problems with previous contraceptive use, mense’s returned after birth, sexual resumption |
| Andualem G et al.,2022  , Ethiopia ^20^ | Cross sectional, Community based study | 402 | Utilization of modern PPFP methods in the extended postpartum period | Maternal education, number of ANC visits, discussed contraceptive use with a partner, previous use of FP, PNC check-up, Linkage to FP unit during child immunization, Attitude towards FP, knowledge towards FP, menses resumption, sexual resumption |
| Gebeyehu NA et al.,2020  , Ethiopia^21^ | Cross sectiona, Facility-based study | 416 | Intention to use contraception | Education, children alive, Attending ANC follow-up, Attending PNC follow-up, Discussion with partner on PPFP, menses resumption, sexual resumption, previous use of contraceptive, Husband approval of contraceptives, knowledge of contraceptive |
| Coomson JL et al.,2019  , Ghana ^22^ | Cross sectional, Facility-based study | 320 | Post partum modern contraceptive use | Age, sexual resumption, menses resumption, ever used modern contraceptive, discussion with partner, partner approval, knowledge about available FP methods |
| Gebremedhin Jnr M et al.,2021  , Ethiopia ^23^ | Cross sectional, Facility-based study | 452 | Immediate PPIUCD Acceptance | Age, residence, parity, Previous FP use, ANC follow-up, prior discussion |
| Geda YF et al.,2021  , Ethiopia ^24^ | Cross sectional, Facility-based study | 286 | Immediate Postpartum intrauterine contraceptive utilization | Age, occupation, educational status, birth interval, discuss about PPFP with partner, needs partner approval to use PPFP, knowledge about PPIUCD |
| Samuel B et al.,2023  , Ethiopia ^25^ | Cross sectional, Facility-based study | 373 | Informed choice of LARC | Education, occupation, husband’s education, previous contraception use, Frequency of listening to a radio, heard about LARCs at community conversation, Seen posters with messages about LARCs in the facility |
| Omona K et al.,2020  , Uganda ^26^ | Cross sectional, Facility-based study | 202 | Utilization of IUCD | Education, religion, occupation, culture allow IUCD use, social interaction, IUCD methods are available in this hospital, knowledge on the method, Is IUCD free of charge |
| Rahayuwati L et al.,2023  , Indonesia ^27^ | Cross sectional, Regional study | 280 | Contraceptive use | Not applicable |
| Shiferaw Y et al.,2023  , Ethiopia ^28^ | Cross sectional, Facility-based study | 392 | Utilization of immediate postpartum intrauterine contraceptive device | Age, birth interval, use of FP in the past, plan to have another child, ever heard of IUCD, attitude towards the use of PPIUCD |
| Belayihun B et al.,2021  , Ethiopia ^29^ | Cross sectional, Facility-based study | 884 | Immediate postpartum LARC use | Age, marital status, education, residence, children alive, ever use FP, PPFP information, ANC follow-up, use of MWHs, Mode of delivery, child immunization |
| Abera Y et al.,2015  , Ethiopia ^30^ | Cross sectional, Community based study | 703 | Contraceptive use | Age, marital status, education, husband’s education, children alive, fertility desire, Postpartum period, menses resumption, place of delivery, ANC visit, LAM knowledge |
| Gejo NG et al.,2019  , Ethiopia ^31^ | Cross sectional, Facility-based study | 368 | Postpartum family planning use | Education, monthly income, menses resumption, sexual resumption, currently breastfeeding, parity, ANC, Problem experienced while using contraception, postpartum period |
| Hounkponou F et al.,2019  , Africa ^32^ | Cross sectional, Community based study | 453 | Postpartum Contraceptive Use | Contraceptive method use history, sexual resumption, Discussion about contraceptive use, partner’s permission needed |
| Tafere TE et al.,2018, Ethiopia ^33^ | Prospective cohort, Facility-based study | 823 | Post-partum modern family planning use | Residence, education, occupation, Counselling on BPRCP, Counselling on breast feeding, at least one PNC visit, Satisfaction on ANC service during pregnancy |
| Jima GH et al.,2020, Ethiopia ^34^ | Cross sectional, Community based study | 1160 | Postpartum family planning utilization | Education, residence, partner’s education, knowledge, attitude, ANC, PNC, Prior use of Contraception, sexual resumption, Couple discussion on FP, Husband approval of FP use, Socio cultural influence |
| Hagos H et al.,2020, Ethiopia ^35^ | Cross sectional, Facility-based study | 182 | Postpartum IUCD utilization | Not applicable |
| Assefaw M et al., 2021, Ethiopia ^36^ | Case control, Facility-based study | 420 | Uptake of a postpartum intra uterine contraceptive device | Age, marital status, education, family size, children alive, parity, Plan to have children, ANC, ever use FP, Ever heard about PPIUCD, Knowledge about PPIUCD, attitude about PPIUCD |
| Usso AA et al., 2021, Ethiopia ^37^ | Cross sectional, Facility-based study | 530 | Utilization of LARC | Residence, monthly income, mode of delivery, time to reach near health facility, previous use of modern contraceptives, ANC, discussion with partner on contraceptives, main decision maker on contraceptives, Attitude toward LARC, Disrespect and abuse during childbirth |
| Tariku M et al., 2022, Ethiopia ^38^ | Cross sectional, Facility-based study | 418 | Uptake of LARC | Age, ethnicity, occupation, planned birth, ANC visits, Heard about LARC, Previous use of LARC, |
| Nigussie AT et al., 2016, Ethiopia ^39^ | Cross sectional, Community based study | 545 | Currently FP utilization | Education, birth interval, complementary feeding, ANC visit, place of delivery, Have PNC visit, discuss partner, husband decide, decide yourself, postpartum period, Information about FP, attitude |
| Ismael K et al., 2023, Ethiopia ^40^ | Cross sectional, Community based study | 804 | Timely initiation of postpartum contraceptive utilization | Education, monthly income, residence, Receive ANC, place of delivery, have PNC follow-up, menses returned, Discussed family planning with husband |
| Joshi AK et al., 2020  , Nepal ^41^ | Cross sectional, Community based study | 427 | Postpartum Family Planning method utilization | Ethnicity, education, occupation, husband’s occupation, husband’s education, menses resumption, knowledge of FP, past FP use, use of media, PNC visit |
| Wayessa MB et al., 2020  , Ethiopia ^42^ | Prospective cohort, Facility based study | 726 | IUCD uptake | Age at first marriage, Gravidity, history of abortion, number of children, children’s sex, sex preference, plans to have more child, decision on having child, level of knowledge, attitudes towards IUCD |
| Tesfaye T et al., 2023, Ethiopia^43^ | Cross-sectional, Facility based study | 470 | Postpartum Family Planning method utilization | Not applicable |
| Mickler AK et al., 2021, Ethiopia ^44^ | Cross-sectional, Facility based study | 936 | Postpartum contraception use | Age, residence, parity, prior use of contraception, delivery mood, delivery attendant, facility type, availability of contraceptive methods |
| Pearson E et al., 2020, Tanzania ^45^ | Cluster-randomized controlled trial, Facility based study | 14950 | IUCD uptake | Age, education, parity, marital status, religion, |
| Abdulkadir Z et al., 2020, Nigeria^46^ | Randomized controlled trial, Facility based study | 150 | Postpartum contraception use | Not applicable |
| Hersh AR et al., 2018, Colombia^47^ | Multicenter randomized-controlled trial, Facility based study | 240 | Long-acting reversal contraception uptake | Not applicable |
| Mruts KB et al., 2022, Ethiopia ^48^ | Cross- sectional, Facility based study | 1650 | Postpartum contraception use | Age, residence, region, quintile, education, employment, marital status, parity, religion, intended last child, sex of last child, ANC, Place of delivery, PNC, fertility intention, family planning massages, distance of health services. |
| Ayiasi RM et al., 2015, Uganda ^49^ | Community based intervention study | 1385 | Postpartum modern contraception use | Initiation of breast feeding, pre- lacteal test, contraceptive methods, user willingness, pregnancy test, contraceptive choose willingness. |
| Karra M et al., 2017, Sri Lanka ^50^ | Cross- sectional, Facility based study | 13731 | IUCD uptake | Not applicable |
| Wu WJ et al., 2020, Nepal ^51^ | Pre–post-intervention study | 953 | Postpartum modern contraception use | Age, monthly expenditure, caste, delivery mode, sex of child |
| Pradhan E et al., 2019, Nepal ^52^ | Cluster randomized controlled trial, Facility based study | 75587 | IUCD uptake | Age, education, Time to travel from home to hospital, parity, ethnicity, region, abortion |
| Kaewkiattikun K., 2017, Thailand ^53^ | Randomized controlled trial, Facility-based study | 233 | Long-acting reversal contraception uptake | Age, education, occupation, income, knowledge about contraception, pregnancy intention, previous family planning, ANC, mode of delivery |
| Rajan S et al., 2016, India ^54^ | Cross- sectional, Regional study | 2733 | Postpartum modern contraception use | Age, age at marriage, education, religion, caste, wealth index, number of living children, city, residence |
| Chhabra HK et al., 2016, India ^55^ | Cross- sectional, Facility-based study | 117 | Postpartum contraception uptake | Not applicable |
| Puri MC et al., 2021, Nepal ^56^ | Cluster randomized control trial, Facility-based study | 21280 | Unmet need of postpartum contraceptive uptake | Age, age at marriage, education, religion, ethnicity, wealth index, residence, abortion, months since delivery, sex composition of living child. |
| Nurcahyani L et al., 2023, Indonesia ^57^ | Quasi-experimental methods, Facility-based study | 110 | Postpartum contraception uptake | Age, parity. |
| Willcox ML et al., 2021, Uganda ^58^ | Focus group discussions, In-depth interviews, Community-based study | 323 | Postpartum contraception uptake | Not applicable |
| Puri MC et al.,  2020, Nepal ^59^ | In-depth interviews, Facility-based study | 25 | Immediate postpartum intrauterine device use | Not applicable |
| Puri MC et al.,  2020, Nepal ^60^ | In-depth interviews, Facility-based study | 24 | Postpartum contraception uptake | Not applicable |
| Jalang’o R et al., 2017, Kenya ^61^ | Mixed method, Facility-based study | 20 | Postpartum family planning uptake | Not applicable |

**Quality assessment of the included studies**

**Supplemental Table 08.** Newcastle-Ottawa scale assessment of study quality for **cross-sectional study**

| Author | Selection | | | |  | Comparability |  | Outcome | | Study quality |
| --- | --- | --- | --- | --- | --- | --- | --- | --- | --- | --- |
|  | 1 | 2 | 3 | 4 |  | 5 |  | 6 | 7 |  |
|  | Representativeness of the sample | Sample size | Ascertainment of exposure | Non-respondents |  | The subjects in different outcome groups are comparable, based on the study design or analysis. Confounding factors are controlled. |  | Assessment of outcome | Statistical test is appropriate |  |
| Arero WD et al., 2022, Ethiopia | * | * | * | * |  | * |  | * | * | 7 |
| Murry LL et al.,2021, India | * | * | * | * |  | * |  | * |  | 6 |
| Adofo E et al.,2021, Ghana | * | * | * | * |  | * |  | * | * | 7 |
| Wudineh KG et al.,2023, Ethiopia | * | * | * | * |  | * |  | * | * | 7 |
| Melkie A et al.,2021, Ethiopia | * |  | * | * |  | * |  | * | * | 6 |
| Kiondo SK et al.,2020, Tanzania | * | * | * | * |  | * |  | * | * | 7 |
| Silesh M et al.,2022, Ethiopia | * | * | * | * |  | * |  | * | * | 7 |
| Nugussa B et al.,2023, Ethiopia | * | * | * | * |  | * |  | * | * | 7 |
| Tilahun T et al.,2022, Ethiopia | * | * | * | * |  | * |  | * | * | 7 |
| Getaneh M et al.,2021, Ethiopia | * | * | * | * |  | * |  | * | * | 7 |
| Agula C et al.,2022, Ghana | * | * | * | * |  | * |  | * | * | 7 |
| Abebe GF et al.,2023, Ethiopia | * | * | * | * |  | * |  | * | * | 7 |
| Ezekiel MJ et al.,2022, Tanzania | * | * | * | * |  | * |  | * | * | 7 |
| Tafa L et al.,2021, Ethiopia | * | * | * | * |  | * |  | * | * | 7 |
| Nakiwunga N et al.,2022, Uganda | * | * | * | * |  | * |  | * | * | 7 |
| Abraha TH et al.,2017, Ethiopia | * | * | * | * |  | * |  | * | * | 7 |
| Andualem G et al.,2022, Ethiopia | * | * | * | * |  | * |  | * | * | 7 |
| Gebeyehu NA et al.,2020, Ethiopia | * | * | * | * |  | * |  | * | * | 7 |
| Coomson JL et al.,2019, Ghana | * | * | * | * |  | * |  | * | * | 7 |
| Gebremedhin Jnr M et al.,2021, Ethiopia | * | * | * | * |  | * |  | * | * | 7 |
| Geda YF et al.,2021, Ethiopia | * | * | * | * |  | * |  | * | * | 7 |
| Samuel B et al.,2023, Ethiopia | * | * | * | * |  | * |  | * | * | 7 |
| Omona K et al.,2020, Uganda | * |  | * | * |  |  |  | * | * | 5 |
| Rahayuwati L et al.,2023, Indonesia | * | * | * |  |  | * |  | * | * | 6 |
| Shiferaw Y et al.,2023, Ethiopia | * | * | * | * |  | * |  | * | * | 7 |
| Belayihun B et al.,2021, Ethiopia | * | * | * | * |  | * |  | * | * | 7 |
| Abera Y et al.,2015  , Ethiopia | * | * | * | * |  | * |  | * | * | 7 |
| Gejo NG et al.,2019  , Ethiopia | * | * | * | * |  | * |  | * | * | 7 |
| Hounkponou F et al.,2019, Africa | * | * | * | * |  | * |  | * | * | 7 |
| Jima GH et al.,2020, Ethiopia | * | * | * | * |  | * |  | * | * | 7 |
| Hagos H et al.,2020, Ethiopia | * | * | * | * |  | * |  | * | * | 7 |
| Usso AA et al., 2021, Ethiopia | * | * | * | * |  | * |  | * | * | 7 |
| Tariku M et al., 2022, Ethiopia | * | * | * | * |  | * |  | * | * | 7 |
| Nigussie AT et al., 2016, Ethiopia | * | * | * | * |  | * |  | * | * | 7 |
| Ismael K et al., 2023, Ethiopia | * | * | * | * |  | * |  | * | * | 7 |
| Joshi AK et al., 2020, Nepal | * | * | * | * |  | * |  | * | * | 7 |

**Supplemental Table 09.** Newcastle-Ottawa scale assessment of study quality for **case-control study**

| Author | Selection | | | |  | Comparability | |  | Exposure | | | Study quality |
| --- | --- | --- | --- | --- | --- | --- | --- | --- | --- | --- | --- | --- |
|  | 1 | 2 | 3 | 4 |  | 5A | 5B |  | 6 | 7 | 8 |  |
|  | Is the case definition adequate? | Representativeness of the cases | Selection of controls | Definition of controls |  | Case-control comparable on basis of age | Case-control comparable on other factor(s) |  | Ascertainment of exposure | Same method of ascertainment for cases and control | Non-response rate |  |
| Hagos KN et al.,2023  Ethiopia | * | * | * | * |  | * | * |  | * | * | * | 9 |
| Assefaw M et al., 2021, Ethiopia | * | * | * |  |  | * | * |  | * | * | * | 8 |

**Supplemental Table 10.** Newcastle-Ottawa scale assessment of study quality for **cohort study**

| Author | Selection | | | |  | Comparability | |  | Outcome | | | Study  quality |
| --- | --- | --- | --- | --- | --- | --- | --- | --- | --- | --- | --- | --- |
|  | 1 | 2 | 3 | 4 |  | 5A | 5B |  | 6 | 7 | 8 |  |
|  | Exposed cohort truly representative | Non-exposed cohort drawn from the same community | Ascertain-ment of exposure | Outcome of interest not present at start |  | Cohorts comparable on basis of age | Cohorts comparable on other factor(s) |  | Quality of outcome assessment | Follow-up long enough for outcomes to occur | Complete accounting for cohorts |  |
| Mruts KB et al.,2023, Ethiopia | * |  | * | * |  |  | * |  | * | * | * | 7 |
| Maeregayehu T et al.,2022, Ethiopia | * |  | * | * |  |  | * |  | * | * | * | 7 |
| Tafere TE et al.,2018, Ethiopia | * |  | * | * |  |  | * |  | * | * | * | 7 |
| Wayessa MB et al., 2020  , Ethiopia | * | * | * | * |  | * | * |  | * | * | * | 9 |

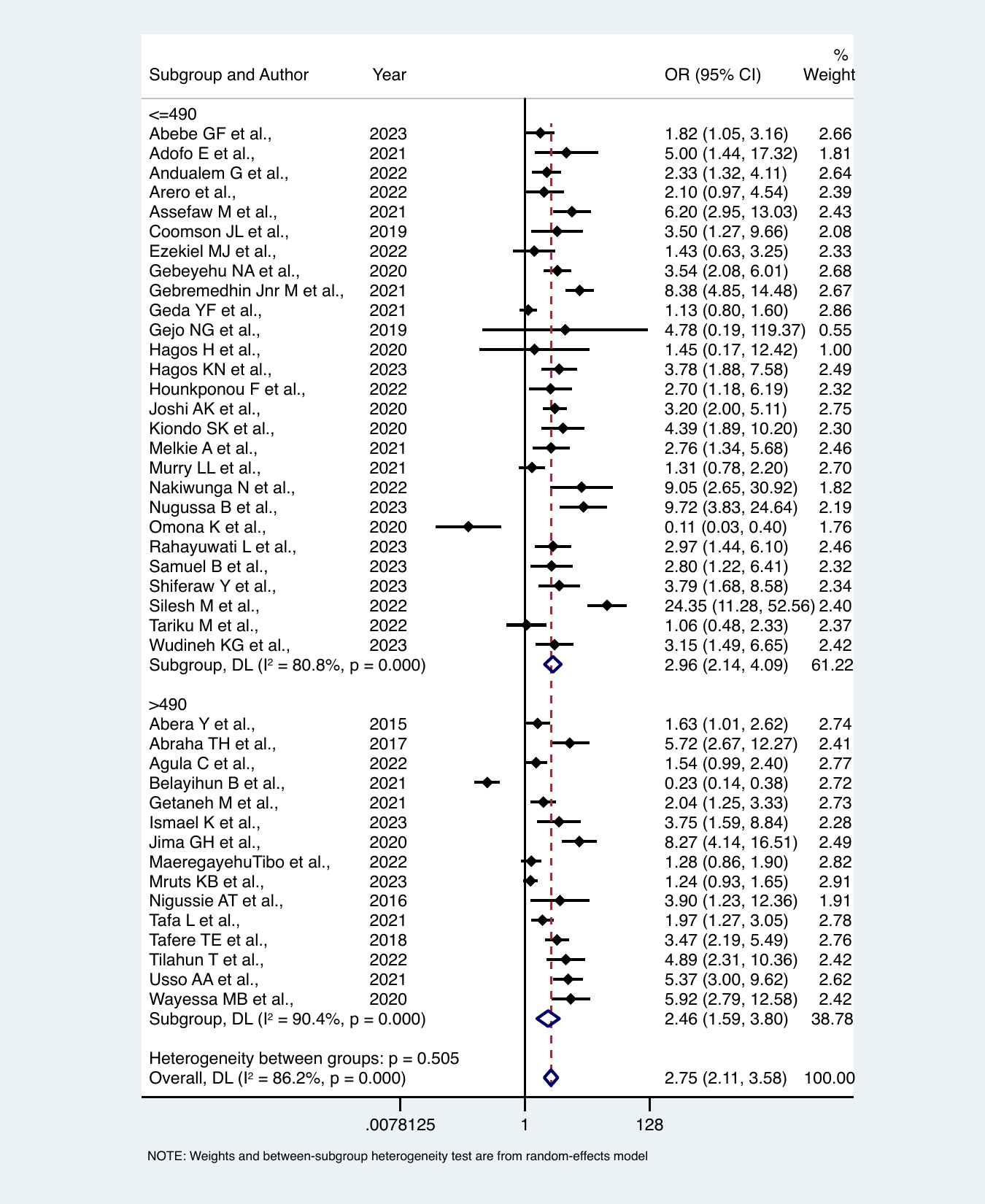

**Supplementary figure 01:** Summary effects of family planning counselling on modern contraception uptake in the post-partum period stratified across sample size

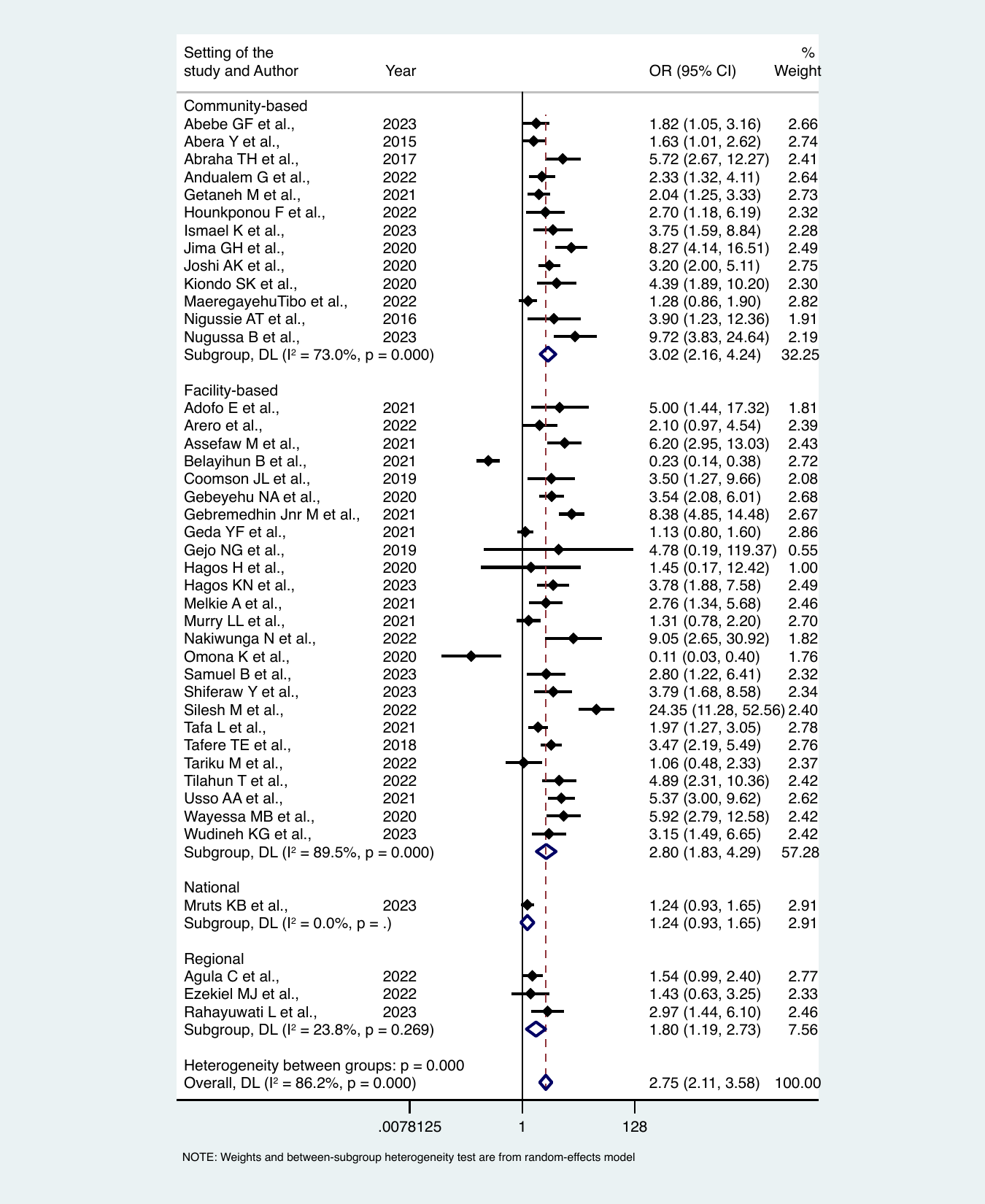

**Supplementary figure 02:** Summary effects of family planning counselling on modern contraception uptake in the post-partum period stratified across study setting

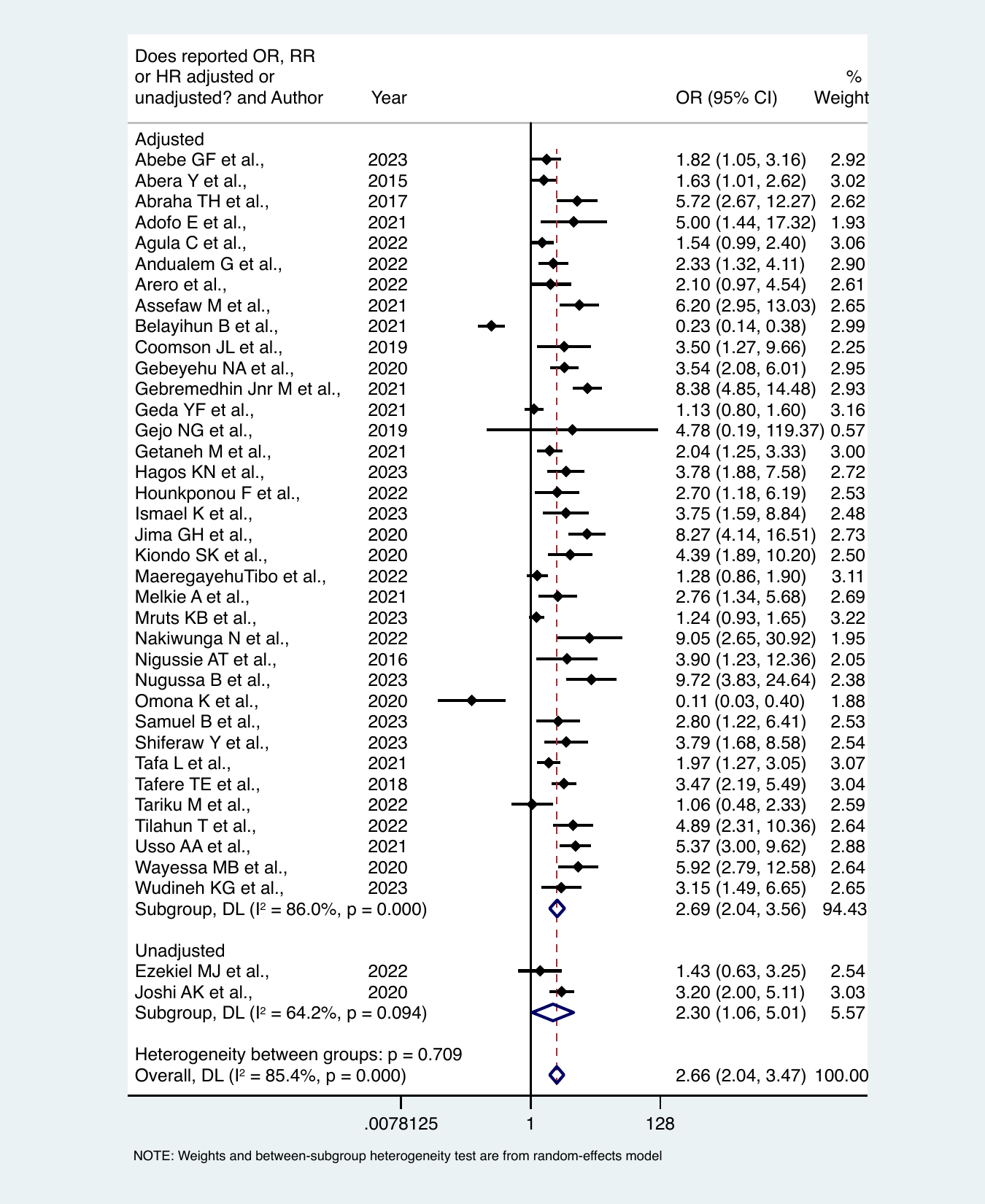

**Supplementary figure 03:** Summary effects of family planning counselling on modern contraception uptake in the post-partum period stratified across confounder adjustment

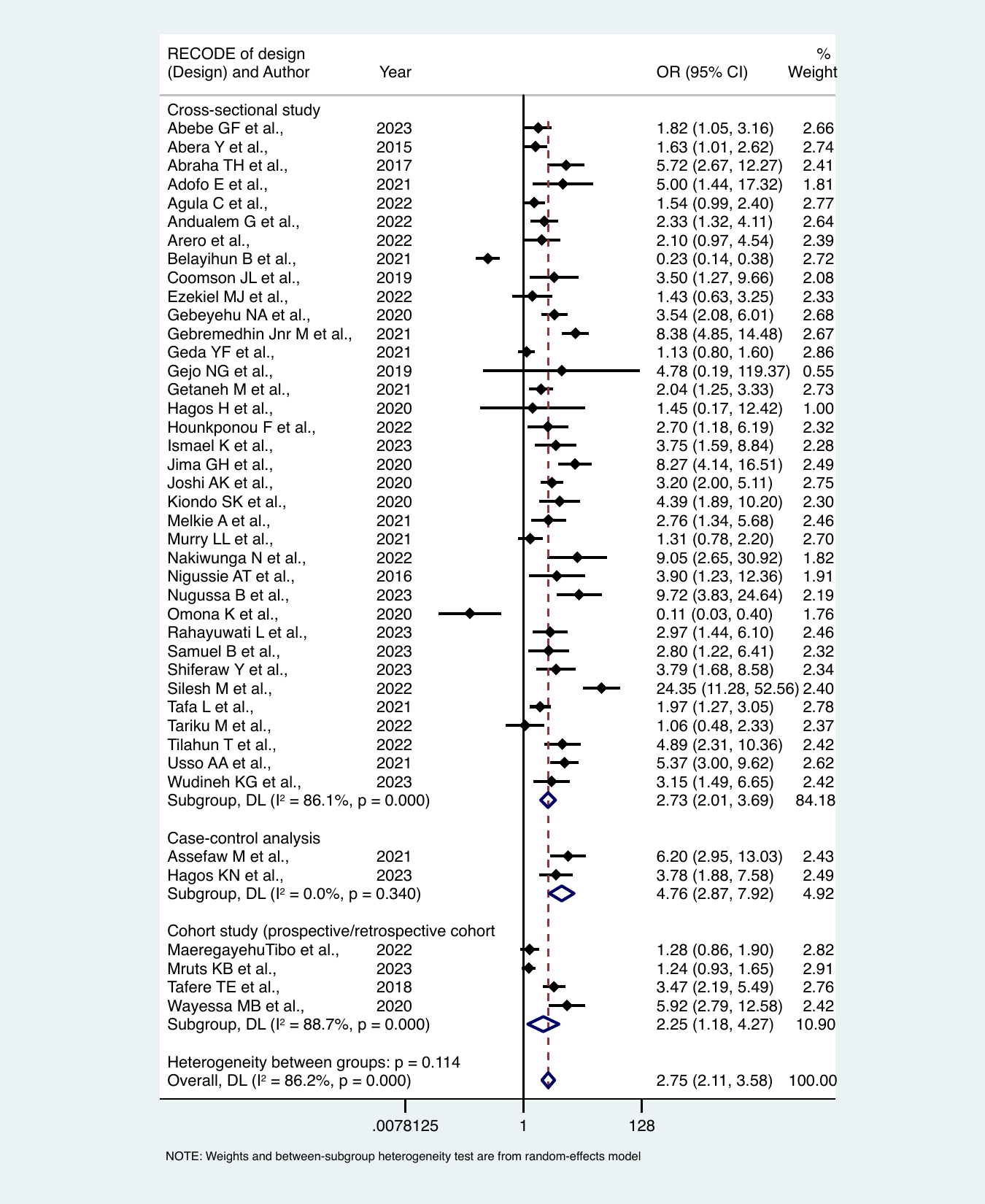

**Supplementary figure 04:** Summary effects of family planning counselling on modern contraception uptake in the post-partum period stratified across study design

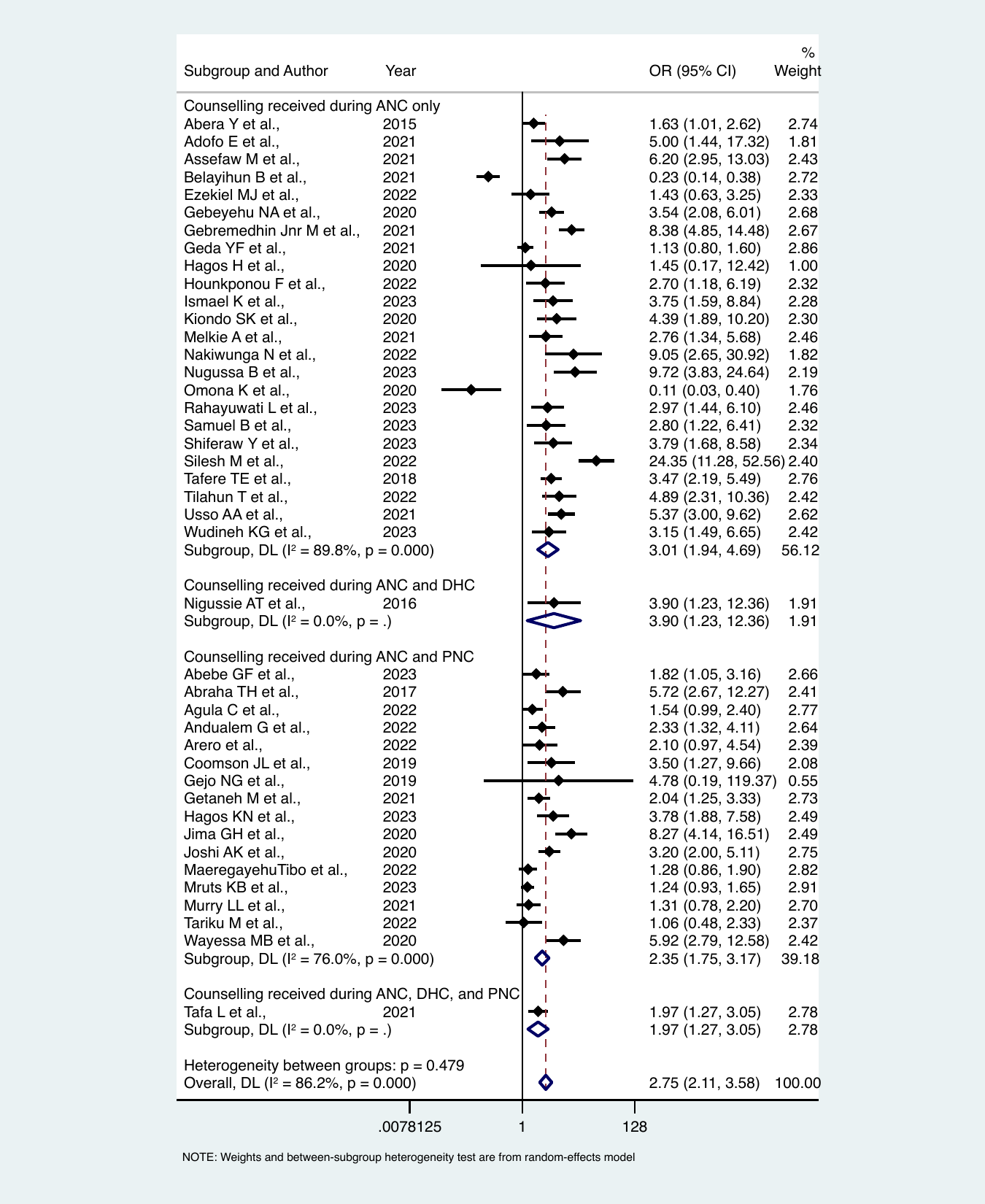

**Supplementary figure 05:** Summary effects of family planning counselling on modern contraception uptake in the post-partum period stratified across stage of maternal healthcare when counselling received

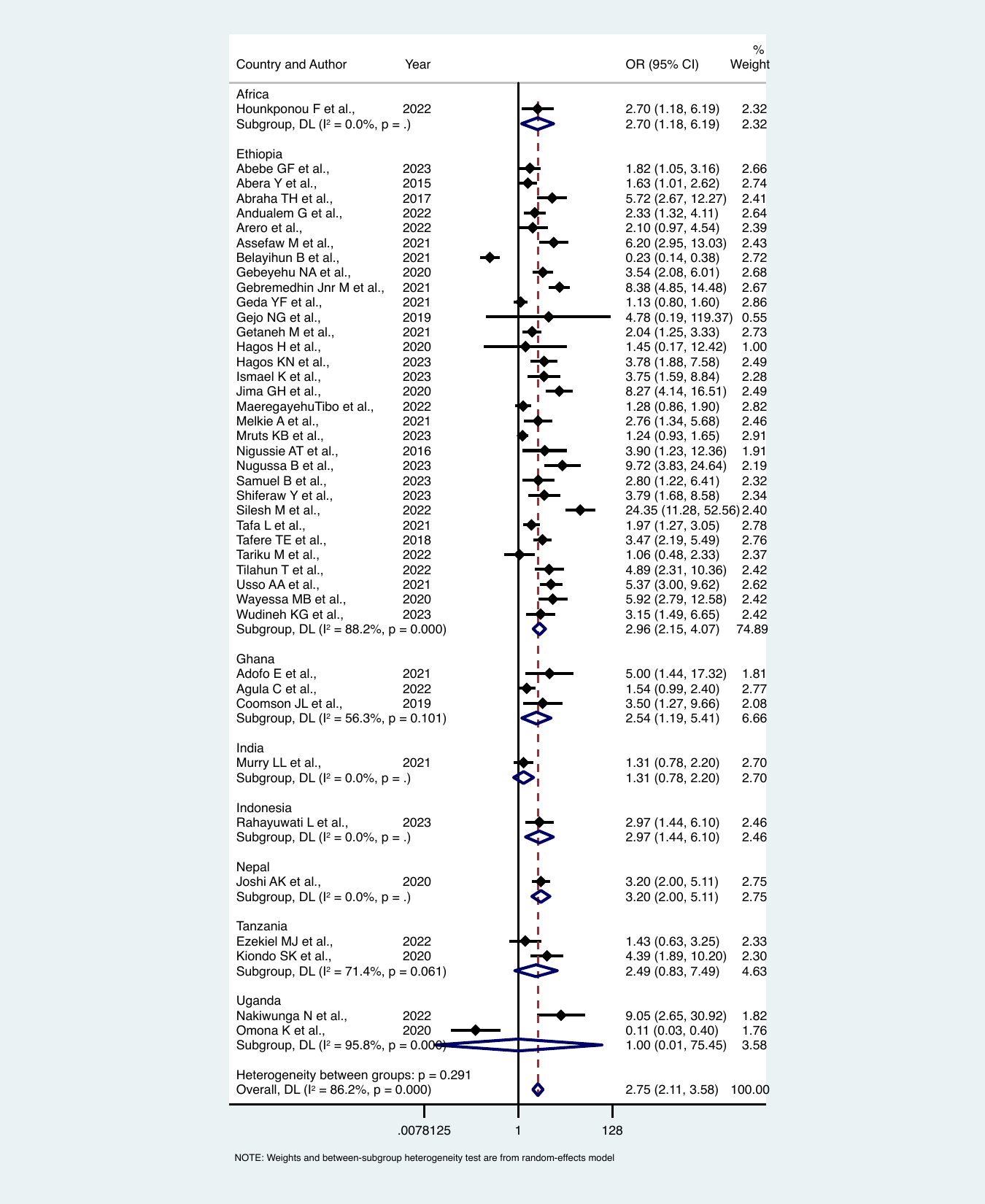

**Supplementary figure 06:** Summary effects of family planning counselling on modern contraception uptake in the post-partum period stratified across countries of included studies.

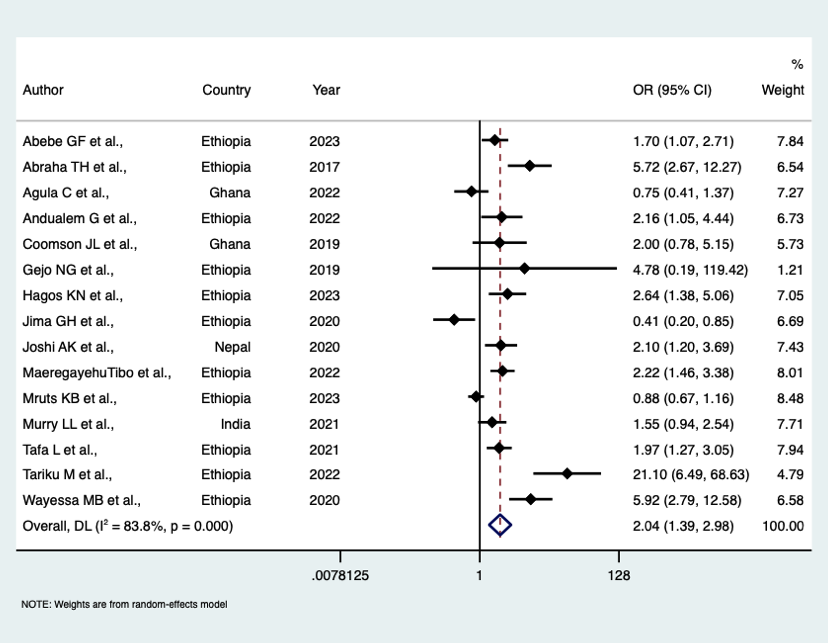

**Supplementary figure 07:** Effects on contraception counselling during accessing postnatal care on modern contraception uptake during accessing postnatal healthcare services.

38.	Tariku M LB, Tantu T,, B D. Uptake of Immediate Postpartum LARCs and Associated

Factors among Mothers Who Gave Birth at Hawassa University

Comprehensive Specialized Hospital, Hawassa, Ethiopia. *International Journal of Reproductive Medicine* 2022; **2022**: 8.

39.	Nigussie AT GDaTG. Postpartum Family Planning Utilization and Associated Factors among Women

who Gave Birth in the Past 12 Months, Kebribeyah Town, Somali Region,

Eastern Ethiopia. *Journal of Women's Health Care* 2016; **6**.

40.	Ismael K CT, Abdo M. Timely initiation of postpartum contraceptive

utilization in Sebata Hawas district, Ethiopia:

A cross-sectional study. PLOS Glob Public Health. *PLOS Glob Public Health* 2023; **3**.

41.	Joshi K A TDP, Poudyal A, Shrestha N, Acharya U, Dhungana G P. Utilization of Family Planning Methods Among Postpartum Mothers in Kailali District, Nepal. *International Journal of Women's Health* 2020; **12**: 487–94.

42.	Wayessa M B ATW, Habtewold E M, Adlo A M, Teklu A M, Abeya S G, Negero W O. Focused Family Planning Counseling Increases

Immediate Postpartum Intrauterine Contraceptive

Device Uptake: A Quasi-Experimental Study. *Journal of Contraception* 2020; **11**(91–102	).
